# Provider Experiences with AI Clinical Decision Support in eSanjeevani

**DOI:** 10.64898/2026.10.12.26365316

**Authors:** Pranshu Bhagat, Shweta Rana, Karuna Nidhi Kaur, Shekar Sivasubramanian, Vipin Garg, Satish Khalikar, Tonmoy Dutta, Ashish Chander, Sanjay Sood, Harpreet Singh

## Abstract

**Background:** Clinical decision support systems (CDSS) are increasingly embedded in national telemedicine services, however how different stakeholders who use them experience these tools, particularly in low-and middle-income countries, is less explored. In the assisted model of India’s eSanjeevani service, spoke health workers enter structured symptoms and hub clinicians review a symptom summary and an artificial intelligence (AI)-generated differential diagnosis.

**Objective:** To describe role-specific experiences of eSanjeevani’s CDSS functions and to estimate how acceptance coexists with specific reported difficulties within each role.

**Methods:** We analyzed a program feedback web survey completed between 11 July and 29 July, 2025 by hub clinicians and spoke providers. Prespecified item profiles covered review of summaries and suggested diagnoses, the perceived relation of suggestions to the clinician’s own diagnosis, ease of use and intention to use (hub), and symptom-capture utility, question quantity, question relevance, summary presentation and satisfaction (spoke). Within-role contrasts were reported as absolute differences in acceptance with Newcombe 95% CIs. Short open comments underwent content analysis.

**Results:** The cohort comprised 190 hub and 662 spoke respondents from 13 and 15 states (21.4% and 19.3% of eligible providers). Among hub respondents, 76.8% regularly reviewed the symptom summary and 58.9% the suggested diagnoses; 72.6% judged suggestions matched (32.6%) or appropriate but different (40.0%), 7.9% mostly inappropriate, and 11.1% reported that suggestions were mostly not generated. Overall, 85.3% intended to keep using the tools. Affirmative intention was 98.4% when suggestions matched, 66.7% when they were mostly inappropriate (difference vs matched or appropriate-23.2 percentage points, 95% CI-48.5 to-4.0) and 61.9% when mostly absent (-25.7, 95% CI-47.1 to-7.3); technical issues, reported by 38.4%, made little difference (-2.8, 95% CI-14.1 to 7.3). Among spoke respondents, 63.4% rated question quantity appropriate, with miscalibration in both directions (15.6% too many; 16.6% too few); 89.9% found the questions relevant and 91.8% found the summary clear. Satisfaction was 75.3% and did not differ materially between those reporting too many questions and those reporting an appropriate number (-2.7, 95% CI-12.8 to 6.0). Comments located spoke difficulties in question burden and missing complaints, and hub difficulties in freezing and in the scope and availability of suggestions. Estimates were stable across cohort, ambiguity and state-exclusion sensitivity analyses.

**Conclusions:** Acceptance of eSanjeevani’s CDSS was high in both roles. For hub clinicians it tracked the perceived quality and availability of AI suggestions rather than technical friction; spoke workers accepted the question burden, but its two-directional miscalibration points toward complaint-specific, adaptive questioning. These provider reports do not measure diagnostic accuracy. They identify log-based and audit-based indicators that the next evaluation cycle should measure directly.

## Introduction

India’s national telemedicine service, eSanjeevani, is one of the largest public telemedicine programs worldwide. By September 15, 2026, it had recorded more than 500 million teleconsultations, delivered through more than 142,000 Ayushman Arogya Mandir spokes, 19,473 hubs and 901 online outpatient departments [1]. Most of this activity runs through an assisted, provider-to-provider model. A community health officer or other primary care provider at a spoke registers the patient, records the presenting complaints and connects to a medical officer or specialist at a hub, who then makes the diagnosis and issues an electronic prescription. Programmatic data from the expansion phase show that this assisted model accounted for more than 93% of national use [2]. The assisted design shifts a substantial share of clinical information gathering to nonphysician spoke workers, and commentators have identified triage, health worker training and the absence of feedback loops as its weak points [3].

Clinical decision support systems (CDSS) are an obvious response to this division of labor, but the evidence that they change care is more modest than their promise. Across 122 controlled trials, computerized CDSS increased the proportion of patients receiving recommended care by only 5.8% (95% CI 4.0%-7.6%), with substantial heterogeneity [4]. Earlier reviews linked success to support delivered automatically within the clinician’s workflow, at the time and place of decision-making [5], and found that evidence on workload, efficiency and unintended effects remained sparse [6]. For decision support delivered on mobile devices in primary care, a Cochrane review identified no trial that reported provider acceptability or satisfaction [7]. Diagnostic suggestion tools add a further layer of uncertainty. Patient-facing symptom checkers listed the correct diagnosis first in only 34% of standardized vignettes [8], and the primary-diagnosis accuracy of digital symptom checkers ranged from 19% to 37.9% across studies [9]. When clinicians see AI suggestions, accurate support can improve their performance while faulty support misleads even experts [10]; in a randomized vignette study, systematically biased model predictions reduced clinician diagnostic accuracy by 11.3 percentage points [11].

Recent work from Kenya makes the gap between acceptance and benefit unusually concrete. A large language model consultation aid embedded in primary care records received positive feedback in 99.5% of the episodes in which clinicians rated it [12], yet physician review found actively harmful recommendations in 7.8% of audited encounters [13], and a pragmatic cluster-randomized trial showed no significant reduction in treatment failure (adjusted odds ratio 0.77, 95% CI 0.55-1.08) [14]. Favorable provider opinion, in other words, is necessary for sustained use but tells us little on its own about whether a deployed tool is working as intended. Deployed models also drift, and widely implemented systems have performed far below expectations once examined locally [15,16]. This is why calls for continual, local monitoring of clinical AI have grown louder [17–19].

eSanjeevani added CDSS functions to its assisted workflow from March 2023. According to the developers’ description, spoke workers complete a structured, branching chief-complaint form, and a gradient-boosted classifier trained on historical consultations uses the structured symptoms, age and sex to display up to three probable diagnoses to the hub clinician. The version deployed from January 2025 is described as covering 300 symptom workflows and 31 diagnoses [20]. The two functions sit at different points in one consultation. A tool that eases the hub clinician’s reasoning may simultaneously add documentation work at the spoke, so a single overall rating of “the CDSS” can conceal very different experiences [21]. Existing provider studies of eSanjeevani have examined telemedicine in general, among doctors nationally [22], community health officers in one region [23] and doctors in Manipur [24], or have compared platform versions at a single hub [25]. A state evaluation in Odisha reported that 94% of community health officers were comfortable with the symptom checker and 90% of hub and sub-hub doctors found the probable-diagnosis feature useful, but it examined these functions within a broader readiness assessment [26]. Clinician surveys of symptom checkers elsewhere describe both streamlined work and new tasks [27] and only modest physician satisfaction [28].

The developers’ implementation report notes that a provider feedback survey was conducted and that most respondents were positive, but no detailed, role-specific results [20]. The present paper is the full report of that survey. We treat it as an implementation study of two role-specific functions rather than as an evaluation of clinical effectiveness, and we read acceptance alongside the specific difficulties that the same respondents reported.

## Objective

The study had three objectives: (1) to describe prespecified, role-specific implementation profiles of eSanjeevani’s CDSS functions among hub clinicians and spoke health workers; (2) to estimate, within each role, how acceptance coexisted with specific reported difficulties, namely perceived inappropriate or absent diagnostic suggestions and technical problems for hub clinicians and perceived question quantity for spoke workers; and (3) to locate reported problems in the workflow using open comments and to translate them into indicators for a future evaluation cycle.

## Methods

### Study Design and Reporting

This was a cross-sectional analysis of a role-specific web survey administered to eSanjeevani providers; the field instruments are reproduced in Multimedia Appendix 1. Reporting follows the STROBE statement for cross-sectional studies [29] and the Checklist for Reporting Results of Internet E-Surveys (CHERRIES) [30]; the completed checklists are provided in Multimedia Appendices 3 and 4, respectively.

### Setting and the Clinical Decision Support Functions

Figure 1 maps the assisted consultation and the points at which the survey took measurements. At the spoke, the provider records chief complaints through a searchable, multilingual symptom list, may add a symptom manually or upload an image, and answers branching multiple-choice questions on attributes such as onset, duration and intensity. The platform then generates a structured symptom summary that travels with the teleconsultation call. At the hub, the clinician can review the summary, modify symptoms and view the suggested differential diagnoses before recording a diagnosis and prescription. The structured entry form is knowledge based; the differential-diagnosis component is a machine-learning model [20]. We relied on the developers’ published description for model details and did not verify which version or functions were active at each site during fieldwork. The survey did not assess diagnostic accuracy.

**Figure 1.**
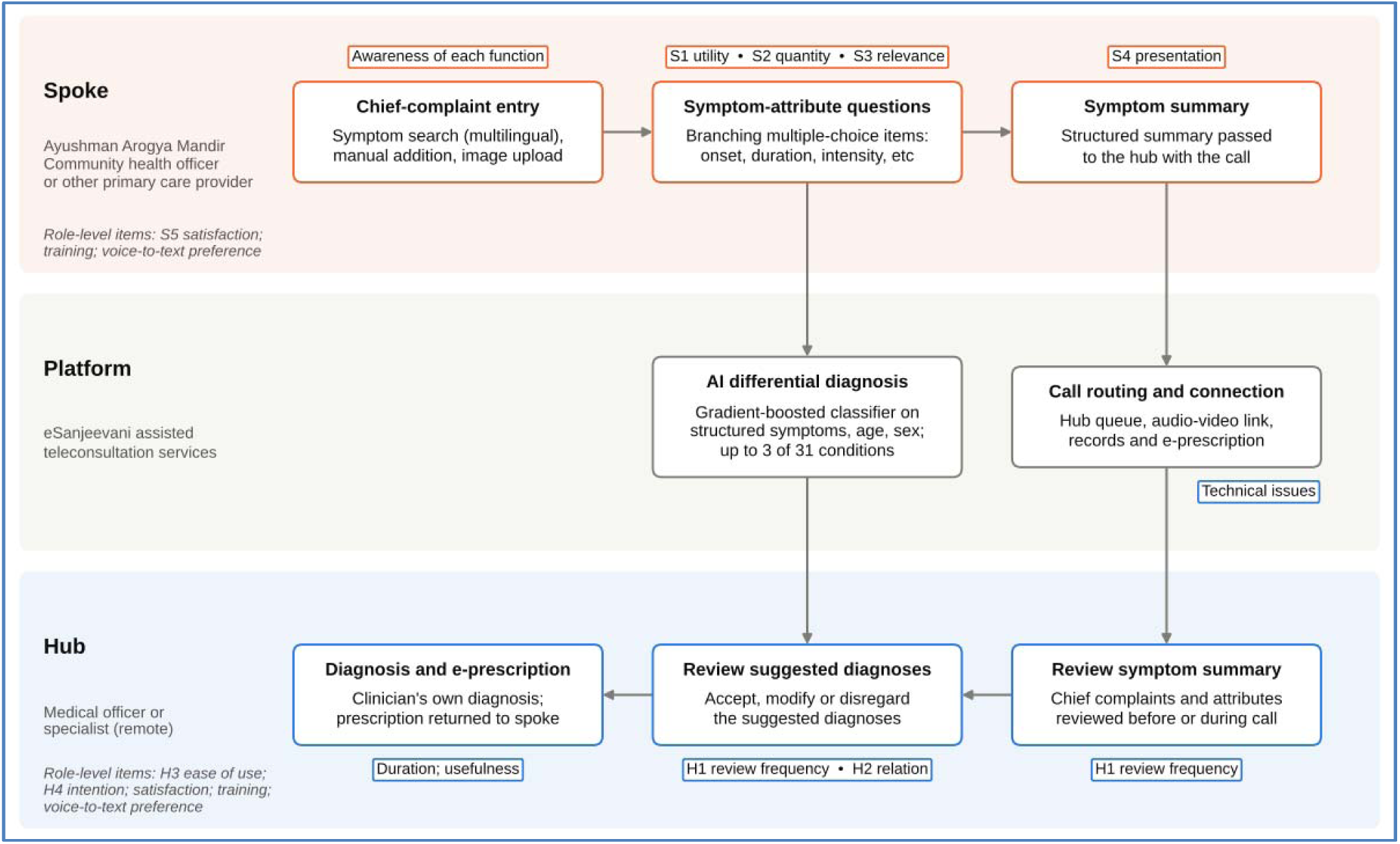
Assisted eSanjeevani consultation workflow, the clinical decision support functions each role encounters and the survey items that measured them. Model details follow the developers’ description of the version deployed from January 2025 [20]. S1-S5 and H1-H4 denote the prespecified spoke and hub profile items; role-level items are listed in italics. Arrows show the intended information flow, not measured respondent-level effects.

### Participants and Recruitment

States and union territories were grouped into six regions; those with fewer than 50,000 cumulative teleconsultations were excluded, and about half of the eligible states in each region were selected with probability proportional to consultation volume. Eligible hub providers were those designated as hub doctors in program records with at least 50 consultations in the preceding 6 months, at least 100 cumulative consultations and at least 6 months on the platform. Eligible spoke providers had at least 75 consultations in the preceding 6 months and at least 100 cumulative consultations. These criteria identified 889 eligible hub providers and 3,426 eligible spoke providers. State program coordinators of the Centre for Development of Advanced Computing (C-DAC) circulated separate hub and spoke survey links through official eSanjeevani communication channels. Participation to the survey was voluntary.

### Survey Instruments

The two forms were designed by the program and research team around the tasks each role performs, drawing on published CDSS usability and acceptance work and on discussion with platform developers. The hub form asked about awareness of the symptom summary, symptom modification and suggested diagnoses; how often the summary and the suggestions were reviewed; how suggestions related to the clinician’s own diagnosis; perceived effect on consultation duration; overall usefulness; checkbox statements on usefulness and ease of use; intention to use the tools regularly; satisfaction; technical problems; training; a voice-to-text preference; and open comments. The spoke form asked about awareness of symptom search, multilingual search, manual addition and image upload; the quantity and relevance of the symptom-attribute questions; presentation of the symptom summary; usefulness; checkbox statements on usefulness and on the chief-complaint section; satisfaction with that section; training; a voice-to-text preference; and open comments. Item wording and response options as exported are reproduced in Tables S1 and S2 in Multimedia Appendix 1, and differences between the pilot and field versions in Table S3 in Multimedia Appendix 1.

The forms were piloted between June 4 to 9, 2025. These records were excluded from the study cohort. The field version was completed between July 11 to 29, 2025.

### Data Processing and Analysis Cohort

Forms recorded the email address of the account used for submission. Before analysis, addresses were normalized and replaced with random study identifiers, and the linkage file was held separately. Age and years of experience were cleaned, qualifications were grouped into role-relevant categories, and state names were harmonized (Table S2 in Multimedia Appendix 2).

### Outcomes

The primary outcome was a prespecified role-specific implementation profile: a fixed set of separately reported item distributions rather than a composite score. For hub clinicians, it comprised frequency of reviewing the symptom summary and the suggested diagnoses (H1), the relation of suggestions to the clinician’s own diagnosis (H2), endorsed ease-of-use statements (H3) and intention to use the tools regularly (H4). For spoke workers, it comprised endorsed symptom-capture statements (S1), perceived quantity (S2) and relevance (S3) of symptom-attribute questions, presentation of the symptom summary (S4) and satisfaction with the chief-complaint section (S5).

Secondary outcomes were within-role coexistence contrasts. For hub clinicians, acceptance was affirmative intention (definitely or probably yes). It was compared between those reporting mostly inappropriate suggestions and those reporting suggestions that matched or were appropriate but different; between those reporting that suggestions were mostly not generated and those whose answer implied that suggestions were available; and between those reporting and not reporting technical problems. For spoke workers, acceptance was satisfaction (satisfied or very satisfied), compared between those reporting too many questions and those reporting an appropriate number. A broader comparator (appropriate or too few) served as a sensitivity analysis. Ambiguous answers and categories outside each comparison were excluded from that contrast only. We also examined full-category cross-tabulations for the two association questions: hub intention by diagnostic-support experience, and spoke satisfaction by question quantity. Operational definitions of every profile item, acceptance measure and contrast are listed in Table S3 in Multimedia Appendix 2.

## Statistical Analysis

Categorical items were summarized as counts and percentages of respondents who answered, with Wilson 95% CIs [31]; checkbox endorsements used to answer respondents as the denominator. For each contrast we report the four cell counts, the joint proportion, the proportion with the problem among those accepting, acceptance in each comparison group and the absolute difference in acceptance (problem group minus comparator) with the Newcombe hybrid score interval [32]. The intervals assume independent respondents and describe sampling variation within the responding cohort; they do not quantify uncertainty about all eSanjeevani providers. For the two association questions, global independence was assessed with a Monte Carlo permutation chi-square test (20,000 permutations), and the two P values were Holm adjusted. As supporting analyses, we fitted one proportional-odds model per role, with intention grouped into three ordered levels (not affirmative, probably yes, definitely yes) or satisfaction grouped into three ordered levels (not favorable, satisfied, very satisfied), the exposure in its original categories and years of experience (<4, 4-10, >10 years) as the only adjustment variable. The proportional-odds assumption was examined by comparing cut-specific binary logistic models. No unasked item was imputed.

Sensitivity analyses were specified for the cohort, ambiguous answers (exclusion, conservative and favorable bounds), role attribution (hub respondents with medical qualifications only), exposure (hub respondents aware of the suggestions and reviewing them at least occasionally), possible misreading of the satisfaction scale (exclusion of very dissatisfied responses from respondents who also rated the tools always useful [33]) and geographic concentration (sequential exclusion of each state with at least five respondents). Analyses used Python 3.13 with pandas 3.0, SciPy 1.18 and statsmodels 0.15.

### Qualitative Content Analysis

Open responses were analyzed with conventional content analysis [34]. Each response was first classified as empty, a placeholder, an explicit “nothing” or “no comment,” or substantive. Substantive responses were grouped into three domains according to the prompt: strengths (what was liked), difficulties (what was liked least or disliked, and descriptions of technical problems) and requested improvements (suggestions and other improvements). A codebook was developed inductively from the full corpus and organized by the function responsible for each problem: CDSS content and logic, interface, platform performance and connectivity, service operations, and training. Coding used keyword rules followed by line-by-line review of every substantive response, with documented overrides wherever the rules misclassified a comment.

Each respondent was counted at most once per theme within a domain; theme frequencies are reported against both the number of respondents commenting in that domain and the whole role cohort. Two prompts listed examples (symptom modification and differential diagnosis for the hub “liked least” item; image uploads, multilanguage support and quicker symptom editing for the improvement items). Responses that only named a listed component, or only repeated a listed example, were tabulated separately so that prompted echoes would not be read as spontaneous reports. Quotations are reproduced verbatim with pseudonymous identifiers.

## Results

### Participants

The export contained 956 submissions (237 hub, 719 spoke). The analysis cohort comprised 190 hub and 662 spoke respondents, representing 842 distinct accounts (Figure 2). These numbers correspond to 21.4% of the 889 eligible hub providers and 19.3% of the 3,426 eligible spoke providers. Because respondents could not be linked to the eligibility list, these figures are approximate participation fractions, not verified response rates.

**Figure 2.**
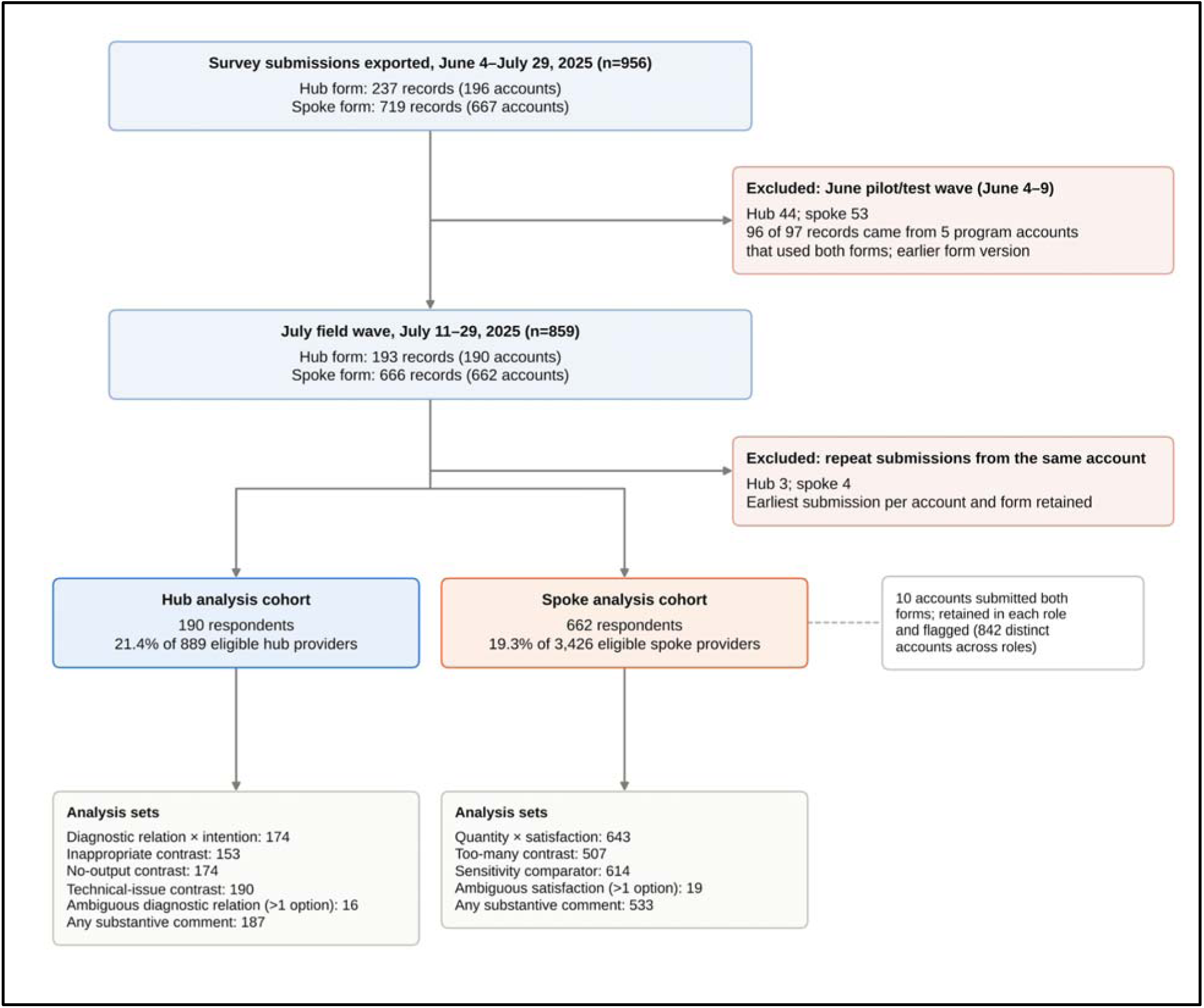
Derivation of the analysis cohort.

The two cohorts differed as their roles would predict (Table 1). Hub respondents had a median age of 33 years (IQR 29-39) and a median of 5 years of experience; 159 of 190 (83.7%) held a medical degree, 8 an AYUSH (Ayurveda, Yoga and Naturopathy, Unani, Siddha and Homoeopathy) degree, and 21 (11.1%) a nursing or community health qualification. Spoke respondents were younger (median 30 years, IQR 27-34); 447 (67.5%) held a nursing degree or diploma and 109 (16.5%) an AYUSH degree. Responses came from 13 states for the hub form and 15 for the spoke form.

**Table 1.** Characteristics of the analysis cohort by role.

| Characteristic | Hub clinicians (n=190) | Spoke health workers (n=662) |
| --- | --- | --- |
| Respondents, n | 190 | 662 |

| Characteristic | Hub clinicians<br>(n=190) | Spoke health workers<br>(n=662) |
| --- | --- | --- |
| **Age (years)** |  |  |
| Median (IQR) | 33 (29-39) | 30 (27-34) |
| <30, n (%) | 52 (27.5) | 321 (49.0) |
| 30-39, n (%) | 95 (50.3) | 290 (44.3) |
| ≥40, n (%) | 42 (22.2) | 44 (6.7) |
| Missing or implausible, n | 1 | 7 |
| **Gender, n (%)** |  |  |
| Female | 85 (44.7) | 385 (58.2) |
| Male | 104 (54.7) | 275 (41.5) |
| Transgender or another gender | 1 (0.5) | 2 (0.3) |
| **Highest qualification, n (%)** |  |  |
| Hub form: MBBS | 114 (60.0) | - |
| Hub form: MBBS with postgraduate qualification | 45 (23.7) | - |
| Hub form: AYUSH degree | 8 (4.2) | - |
| Hub form: Nursing or community health | 21 (11.1) | - |
| Hub form: Other or unclear | 2 (1.1) | - |
| Spoke form: Nursing degree | - | 374 (56.5) |
| Spoke form: Nursing diploma (GNM) | - | 73 (11.0) |
| Spoke form: AYUSH degree | - | 109 (16.5) |
| Spoke form: Community health qualification | - | 44 (6.6) |
| Spoke form: MBBS or higher medical | - | 38 (5.7) |
| Spoke form: Other or unclear | - | 24 (3.6) |
| **Years of experience in health care** |  |  |
| Median (IQR) | 5 (3-11) | 4 (3-7) |
| <4, n (%) | 56 (29.6) | 257 (39.1) |
| 4-10, n (%) | 84 (44.4) | 318 (48.4) |
| >10, n (%) | 49 (25.9) | 82 (12.5) |
| Missing or implausible, n | 1 | 5 |
| **Geography** |  |  |
| States represented, n | 13 | 15 |
| Districts represented, n | 115 | 169 |
| Largest state 1, n (%) | Gujarat 50 (26.3) | Maharashtra 178 (26.9) |
| Largest state 2, n (%) | Andhra Pradesh 50 (26.3) | Jharkhand 100 (15.1) |
| Largest state 3, n (%) | Maharashtra 25 (13.2) | Telangana 98 (14.8) |

### Role-Specific Implementation Profiles

Table 2 and Figure 3 present the prespecified profiles. Hub clinicians engaged more consistently with the symptom summary than with the suggested diagnoses: 76.8% (146/190; 95% CI 70.3%-82.3%) reviewed the summary regularly, compared with 58.9% (112/190; 95% CI 51.8%-65.7%) for the suggestions. Asked how the suggestions related to their own diagnosis, 32.6% (62/190) said they always matched and 40.0% (76/190) that they were mostly clinically appropriate but different. Smaller groups reported mostly inappropriate suggestions, 7.9% (15/190; 95% CI 4.8%-12.6%), or that suggestions were mostly not generated, 11.1% (21/190; 95% CI 7.3%-16.3%); 8.4% (16/190) ticked more than one option (Table S5 in Multimedia Appendix 2). The ease-of-use statements most often endorsed were overall user-friendliness (55.3%) and ease of learning (52.6%); fewer endorsed that the tools worked across different patient cases (32.6%). Intention was high: 85.3% (162/190; 95% CI 79.5%-89.6%) planned to use the tools regularly, including 50.0% (95/190) who answered definitely yes.

**Figure 3.**
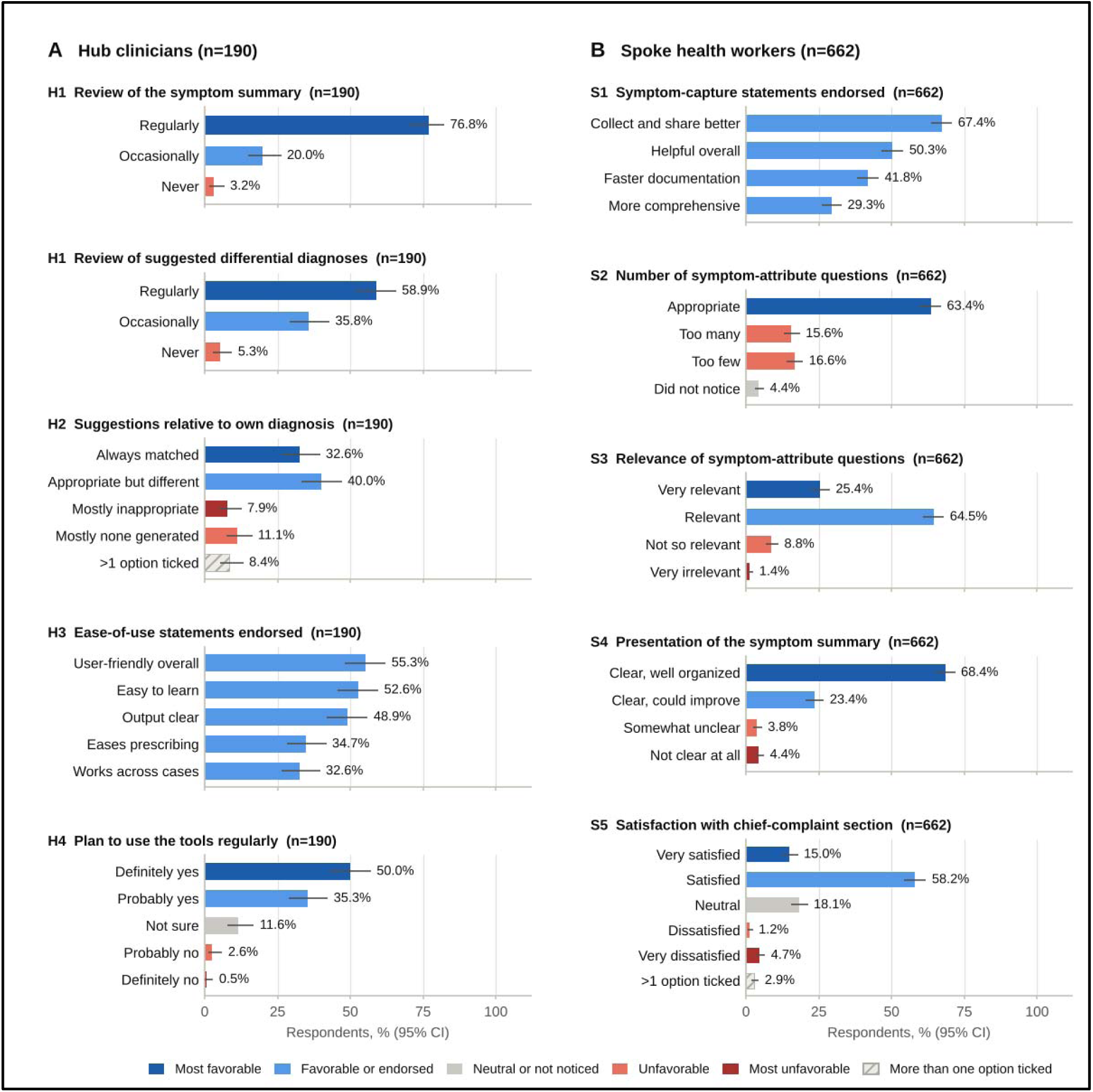
Role-specific implementation profiles. Bars show the percentage of respondents choosing each response (single-response items) or endorsing each statement (checkbox items), with Wilson 95% CIs. Shading indicates favorability, and response labels identify each bar. (A) Hub clinicians (n=190). (B) Spoke health workers (n=662).

**Table 2.** Prespecified role-specific implementation profiles (primary outcome).

| Item and response | n (%) | 95% CI |
| --- | --- | --- |
| **Hub clinicians (n=190)** |  |  |
| **H1 Review of the symptom summary** |  |  |
| Regularly | 146 (76.8) | 70.3-82.3 |
| Occasionally | 38 (20.0) | 14.9-26.3 |
| Never | 6 (3.2) | 1.5-6.7 |
| **H1 Review of suggested differential diagnoses** |  |  |
| Regularly | 112 (58.9) | 51.8-65.7 |
| Occasionally | 68 (35.8) | 29.3-42.8 |
| Never | 10 (5.3) | 2.9-9.4 |
| **H2 Relation of suggestions to own diagnosis** |  |  |
| Always matched my diagnosis | 62 (32.6) | 26.4-39.6 |
| Mostly clinically appropriate but did not match | 76 (40.0) | 33.3-47.1 |
| Mostly clinically inappropriate | 15 (7.9) | 4.8-12.6 |
| Mostly no differential diagnosis was generated | 21 (11.1) | 7.3-16.3 |
| More than one option ticked | 16 (8.4) | 5.2-13.2 |
| **H3 Ease-of-use statements endorsed (checkbox)** |  |  |
| Overall, the CDSS tools are user-friendly | 105 (55.3) | 48.2-62.2 |
| Easy to learn | 100 (52.6) | 45.6-59.6 |
| Information generated is clear and easy to understand | 93 (48.9) | 41.9-56.0 |
| Supports easy and efficient documentation of prescriptions | 66 (34.7) | 28.3-41.7 |
| Functions effectively across different patient cases | 62 (32.6) | 26.4-39.6 |
| **H4 Plan to use the tools regularly** |  |  |
| Definitely yes | 95 (50.0) | 43.0-57.0 |
| Probably yes | 67 (35.3) | 28.8-42.3 |
| Not sure | 22 (11.6) | 7.8-16.9 |
| Probably no | 5 (2.6) | 1.1-6.0 |
| Definitely no | 1 (0.5) | 0.1-2.9 |
| **Spoke health workers (n=662)** |  |  |
| **S1 Symptom-capture statements endorsed (checkbox)** |  |  |
| Helps collect and share symptoms more effectively | 446 (67.4) | 63.7-70.8 |

| **Item and response** | **n (%)** | **95% CI** |
| --- | --- | --- |
| Overall, a helpful tool for remote consultations | 333 (50.3) | 46.5-54.1 |
| Documents symptoms more quickly than manual methods | 277 (41.8) | 38.1-45.6 |
| Captures symptoms and attributes more comprehensively | 194 (29.3) | 26.0-32.9 |
| ***S2 Quantity of symptom-attribute questions*** |  |  |
| Appropriate | 420 (63.4) | 59.7-67.0 |
| Too many | 103 (15.6) | 13.0-18.5 |
| Too few | 110 (16.6) | 14.0-19.6 |
| Did not notice | 29 (4.4) | 3.1-6.2 |
| ***S3 Relevance of symptom-attribute questions*** |  |  |
| Very relevant | 168 (25.4) | 22.2-28.8 |
| Relevant | 427 (64.5) | 60.8-68.1 |
| Not so relevant | 58 (8.8) | 6.8-11.2 |
| Very irrelevant | 9 (1.4) | 0.7-2.6 |
| ***S4 Presentation of the symptom summary*** |  |  |
| Clear and well-organized | 453 (68.4) | 64.8-71.9 |
| Clear but could be improved | 155 (23.4) | 20.3-26.8 |
| Somewhat unclear | 25 (3.8) | 2.6-5.5 |
| Not clear at all | 29 (4.4) | 3.1-6.2 |
| ***S5 Satisfaction with the chief-complaint section*** |  |  |
| Very satisfied | 99 (15.0) | 12.4-17.9 |
| Satisfied | 385 (58.2) | 54.4-61.9 |
| Neutral | 120 (18.1) | 15.4-21.2 |
| Dissatisfied | 8 (1.2) | 0.6-2.4 |
| Very dissatisfied | 31 (4.7) | 3.3-6.6 |
| More than one option ticked | 19 (2.9) | 1.8-4.4 |

For spoke workers, the most frequently endorsed symptom-capture statement was that the chief-complaint section helped them collect and share symptoms more effectively (67.4%); fewer endorsed faster documentation (41.8%) or more comprehensive capture (29.3%). The quantity of symptom-attribute questions was rated appropriate by 63.4% (420/662; 95% CI 59.7%-67.0%). The remainder split almost evenly between too many, 15.6% (103/662; 95% CI 13.0%-18.5%), and too few, 16.6% (110/662; 95% CI 14.0%-19.6%), with 4.4% (29/662) not having noticed. Questions were judged relevant or very relevant by 89.9% (595/662; 95% CI 87.3%-92.0%), and the symptom summary was judged clear by 91.8% (608/662; 95% CI 89.5%-93.7%), although 23.4% added that it could be improved. Among the 643 spoke respondents with an unambiguous satisfaction answer, 75.3% (484/643; 95% CI 71.8%-78.5%) were satisfied or very satisfied.

Supporting items (Tables S6 and S7 in Multimedia Appendix 2) showed near-universal awareness of the summary and suggestions among hub respondents (94.7% and 93.2%) and of symptom search among spoke respondents (94.0%), but lower spoke awareness of multilingual search, 64.4% (426/662; 95% CI 60.6%-67.9%), and image upload, 78.2% (516/660). Of hub respondents giving an unambiguous answer, 67.0% (124/185) perceived consultations as shorter with the tools and 8.6% (16/185) as longer. Prior training on the tools was reported by 38.4% (73/190; 95% CI 31.8%-45.5%) of hub and 48.2% (319/662; 95% CI 44.4%-52.0%) of spoke respondents, whereas 66.3% and 74.6%, respectively, said additional training would help; video tutorials were the most preferred format in both roles.

### Acceptance Alongside Reported Difficulties

Acceptance and reported problems coexisted in both roles, but not uniformly (Table 3 and Figure 4). Technical problems were common, reported by 38.4% (73/190; 95% CI 31.8%-45.5%) of hub respondents, and 61 of 190 (32.1%) reported a technical problem while also intending to keep using the tools. Intention was nearly identical with and without technical problems (83.6% vs 86.3%; difference-2.8 percentage points, 95% CI-14.1 to 7.3). The quality and availability of suggestions told a different story. Affirmative intention was 98.4% (61/62) among respondents whose suggestions always matched, 82.9% (63/76) when suggestions were appropriate but different, 66.7% (10/15) when they were mostly inappropriate and 61.9% (13/21) when they were mostly not generated. Compared with respondents reporting matched or appropriate suggestions, intention was lower among those reporting mostly inappropriate suggestions (-23.2 percentage points, 95% CI-48.5 to-4.0) and among those reporting absent suggestions compared with all respondents whose answers implied that suggestions were available (-25.7 percentage points, 95% CI-47.1 to-7.3). The groups reporting these problems were small, and the intervals are correspondingly wide.

**Figure 4.**
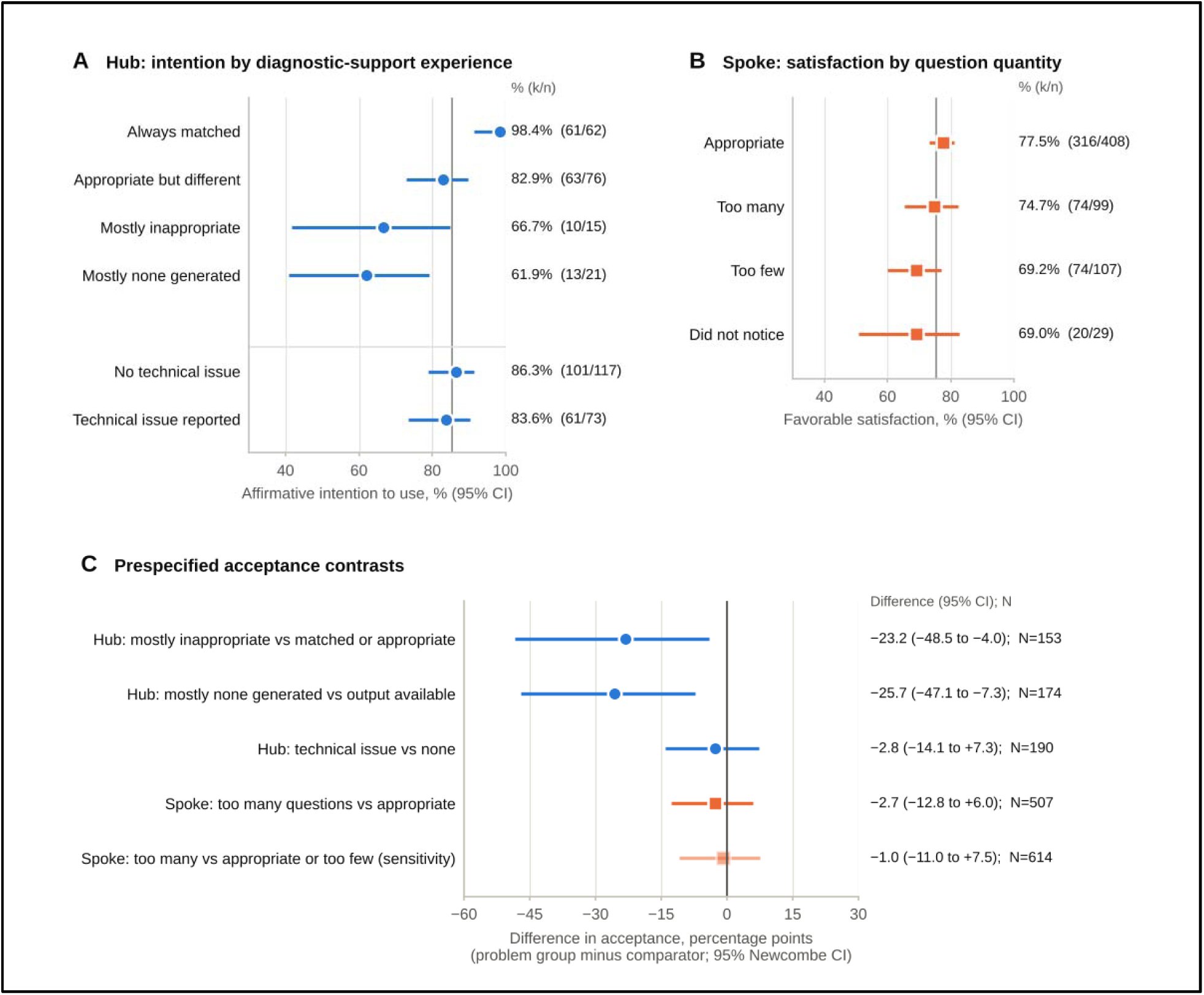
Acceptance by reported experience. (A) Percentage of hub clinicians intending to use the tools regularly (definitely or probably yes), by perceived relation of suggestions to their own diagnosis and by report of technical problems. (B) Percentage of spoke workers satisfied or very satisfied with the chief-complaint section, by perceived quantity of symptom-attribute questions (unambiguous satisfaction answers). Vertical lines mark the overall percentage in each role. (C) Absolute differences in acceptance for the prespecified contrasts, with Newcombe 95% CIs; the lighter marker is the sensitivity comparator. Circles denote hub and squares spoke contrasts. Intervals assume independent respondents.

**Table 3.** Coexistence of acceptance and specific reported difficulties within each role.

| Role | Problem group | Comparator | N | a/b/c/d | Joint, % | Problem among accepting, % | Acceptance with problem, % (95% CI) | Acceptance in comparator, % (95% CI) | Difference, pp (95% CI) |
| --- | --- | --- | --- | --- | --- | --- | --- | --- | --- |
| Hub | Mostly inappropriate suggestions | Matched or appropriate but different | 153 | 10/5/124/14 | 6.5 | 7.5 | 66.7 (41.7-84.8) | 89.9 (83.7-93.9) | -23.2 (-48.5 to -4.0) |
| Hub | Suggestions mostly not generated | Any answer implying suggestions were available | 174 | 13/8/134/19 | 7.5 | 8.8 | 61.9 (40.9-79.2) | 87.6 (81.4-91.9) | -25.7 (-47.1 to -7.3) |
| Hub | Any technical problem | No technical problem | 190 | 61/12/101/16 | 32.1 | 37.7 | 83.6 (73.4-90.3) | 86.3 (78.9-91.4) | -2.8 (-14.1 to 7.3) |
| Spoke | Too many questions | Appropriate number | 507 | 74/25/316/92 | 14.6 | 19.0 | 74.7 (65.4-82.3) | 77.5 (73.2-81.2) | -2.7 (-12.8 to 6.0) |
| Spoke | Too many questions (sensitivity) | Appropriate or too few | 614 | 74/25/390/125 | 12.1 | 15.9 | 74.7 (65.4-82.3) | 75.7 (71.8-79.2) | -1.0 (-11.0 to 7.5) |

Acceptance was affirmative intention to use the tools regularly (definitely or probably yes) for hub clinicians and satisfaction with the chief-complaint section (satisfied or very satisfied) for spoke workers. Cells a/b/c/d are: problem and accepting / problem and not accepting / comparator and accepting / comparator and not accepting. Joint % = a/N; problem among accepting % = a/(a+c). Each contrast excludes ambiguous answers and categories outside its comparison, so N differs by row. Differences are in percentage points (problem group minus comparator) with Newcombe hybrid score 95% CIs; proportions have Wilson 95% CIs. Intervals assume independent respondents. A negative difference indicates lower acceptance in the problem group; it does not imply that the problem caused lower acceptance.

Among spoke workers, 74 of 662 (11.2%) reported too many questions and were nonetheless satisfied. Satisfaction was 74.7% among those reporting too many questions and 77.5% among those reporting an appropriate number (-2.7 percentage points, 95% CI - 12.8 to 6.0); with appropriate or too few as the comparator, the difference was-1.0 percentage points, 95% CI-11.0 to 7.5. Across all four quantity categories, satisfaction ranged from 69.0% to 77.5%.

The full-category tables (Tables S8 and S9 in Multimedia Appendix 2) supported the same reading. Hub intention varied with diagnostic-support experience (permutation chi-square 35.8, n=174; Holm-adjusted P=.002), whereas spoke satisfaction did not vary detectably with perceived question quantity (chi-square 9.81, n=643; Holm-adjusted P=.62). In the supporting proportional-odds models adjusted for experience (Tables S10 and S11 in Multimedia Appendix 2), the odds of higher intention were lower when suggestions were appropriate but different (odds ratio [OR] 0.23, 95% CI 0.11-0.46), mostly inappropriate (OR 0.10, 95% CI 0.03-0.33) or mostly absent (OR 0.12, 95% CI 0.04-0.32) than when they matched; for spoke workers, too many questions was not associated with satisfaction (OR 0.93, 95% CI 0.60-1.46). Cut-specific models gave lower hub odds ratios at the threshold between nonaffirmative and affirmative intention than at the threshold for definitely yes, with wide intervals, so these odds ratios should be read as averages across thresholds (Table S12 in Multimedia Appendix 2).

### Sensitivity Analyses and Measurement Checks

The main estimates changed little across alternative cohorts (Tables S13 and S14 in Multimedia Appendix 2). Hub affirmative intention was 85.0% and spoke satisfaction was 75.1%. Leaving out one state at a time kept these estimates within 83.6%-86.2% and 74.1%-77.0%, respectively. The spoke contrast for too many questions stayed between-4.6 and 1.5 percentage points, and the hub technical-problem contrast between-5.4 and 0.3. Bounding ambiguous spoke satisfaction answers gave 73.6% to 75.8%, and assigning ambiguous hub answers to the problem categories raised the shares reporting mostly inappropriate and mostly absent suggestions to at most 9.5% and 13.2%. Restricting the hub cohort to medically qualified respondents (n=167) gave affirmative intention of 86.2%.

Restricted to respondents who were aware of the suggestions and reviewed them at least occasionally (n=175), the absent-suggestion contrast persisted (-30.3, 95% CI - 54.9 to-9.3), whereas the inappropriate-suggestion contrast weakened (-16.7, 95% CI - 45.2 to 0.4). Second, the satisfaction items may have been misread by some respondents: 8 of 9 hub and 24 of 31 spoke respondents who were very dissatisfied also rated the tools always useful, and few of their comments described problems. Excluding these 24 spoke responses raised satisfaction to 78.2% without changing the quantity contrast (-3.9, 95% CI-14.0 to 4.6).

### Open Comments

At least one substantive comment was provided by 187 of 190 hub and 533 of 662 spoke respondents (Table 4). Hub strengths centered on diagnostic and decision support (63/176 commenting respondents, 35.8%) and ease of use (48/176 commenting respondents, 27.3%); one clinician wrote that the tool would “lead to specific diagnosis and make think about more differential diagnosis”. Spoke strengths centered on structured and comprehensive symptom capture (234/470 commenting respondents, 49.8%), for example that onset and duration options were “already written so no need to type manually”.

**Table 4.** Themes in open comments by role and domain.

| Domain and theme | Respondents, n (% of commenters; % of cohort) | Illustrative quotation |
| --- | --- | --- |
| Hub clinicians (n=190; any substantive comment 187) |  |  |
| *Strengths (respondents commenting: 176)* |  |  |
| Diagnostic and decision support | 63 (35.8; 33.2) | "lead to specific diagnosis and make think about more differntial diagnosis." (P0819) |
| Ease of use | 48 (27.3; 25.3) |  |
| Structured and comprehensive symptom capture | 37 (21.0; 19.5) |  |
| Time saving and speed | 23 (13.1; 12.1) |  |
| Clear summary and presentation | 10 (5.7; 5.3) | "They summerize the case in short, easily readable form..." (P0008) |
| *Difficulties (respondents commenting: 166)* |  |  |
| Technical performance (freezing, lag, errors) | 44 (26.5; 23.2) | "Freezing while symptom modifications, incorrect display" (P0277) |
| Quality or scope of diagnostic suggestions | 22 (13.3; 11.6) | "Diagnosis options are not diagnoses, rather symptoms." (P0114) |
| Interface, navigation and display | 19 (11.4; 10.0) |  |
| Question burden and time to complete | 14 (8.4; 7.4) |  |
| Diagnostic suggestions not generated or not displayed | 14 (8.4; 7.4) | "Freezing, diagnosis not generating timely especially when the network is lslow..." (P0725) |
| Relevance and specificity of questions or suggestions | 13 (7.8; 6.8) |  |
| Prescription, investigation and record functions | 12 (7.2; 6.3) |  |
| Connectivity, audio-video and call handling | 11 (6.6; 5.8) |  |
| Clinician availability, waiting and routing | 9 (5.4; 4.7) |  |
| *Improvements (respondents commenting: 166)* |  |  |
| Image upload and quality (prompted example) | 72 (43.4; 37.9) |  |
| Symptom modification and editing (prompted example) | 35 (21.1; 18.4) |  |
| Prescription, investigation and record functions | 21 (12.7; 11.1) |  |
| Language support (prompted example) | 20 (12.0; 10.5) |  |
| Technical performance (freezing, lag, errors) | 14 (8.4; 7.4) |  |
| Interface, navigation and display | 12 (7.2; 6.3) |  |
| Clinician availability, waiting and routing | 10 (6.0; 5.3) |  |
| Quality or scope of diagnostic suggestions | 9 (5.4; 4.7) |  |
| Spoke health workers (n=662; any substantive comment 533) |  |  |
| *Strengths (respondents commenting: 470)* |  |  |
| Structured and comprehensive symptom capture | 234 (49.8; 35.3) | "Many diseases, onset time, etc. are already written so no need to type manually. Time saving" (P0062) |
| Ease of use | 66 (14.0; 10.0) |  |
| Diagnostic and decision support | 49 (10.4; 7.4) |  |
| Clear summary and presentation | 34 (7.2; 5.1) |  |
| Time saving and speed | 23 (4.9; 3.5) |  |
| *Difficulties (respondents commenting: 268)* |  |  |
| Question burden and time to complete | 82 (30.6; 12.4) | "We can't skip by our choice we have to answer each and each question" (P0649) |
| Symptom or condition coverage gaps | 68 (25.4; 10.3) | "Some basic chief complains are not included in list like skin itching, boils etc" (P0735) |
| Relevance and specificity of questions or suggestions | 28 (10.4; 4.2) | "Asking too many irrelevant question for some complaints" (P0788) |
| Technical performance (freezing, lag, errors) | 10 (3.7; 1.5) |  |
| Interface, navigation and display | 6 (2.2; 0.9) |  |
| *Improvements (respondents commenting: 320)* |  |  |
| Symptom modification and editing (prompted example) | 73 (22.8; 11.0) |  |
| Image upload and quality (prompted example) | 52 (16.2; 7.9) |  |
| Language support (prompted example) | 39 (12.2; 5.9) |  |
| Clinician availability, waiting and routing | 39 (12.2; 5.9) | "Kindly ensure availability of doctors during peak consultation hours." (P0434) |

| Domain and theme | Respondents,<br>n (% of<br>commenters; % of<br>cohort) | Illustrative quotation |
| --- | --- | --- |
| Symptom or condition coverage gaps | 30 (9.4; 4.5) |  |
| Question burden and time to complete | 26 (8.1; 3.9) |  |

Values are the number of respondents with at least one comment coded to the theme, with percentages of respondents commenting in that domain and of the whole role cohort. A respondent could contribute to several themes. Strengths came from the “liked” prompt; difficulties from the “liked least” or “disliked” prompts and, for hub clinicians, descriptions of technical problems; improvements from the suggestion and “other improvements” prompts. The improvement prompt listed image uploads, multilanguage support and quicker symptom editing as examples; 107 of 148 hub and 148 of 206 spoke improvement responses contained only those examples. A further 82 hub respondents only named a component when asked what they liked least and are not counted as difficulties. Quotations are verbatim, including spelling, with pseudonymous identifiers. Full codebook and counts are provided in Tables S16 and S17 in Multimedia Appendix 2.

The difficulties were role specific. Spoke difficulties concerned question burden and time to complete (82/268 commenting respondents, 30.6%), missing symptoms or conditions (68/268 commenting respondents, 25.4%) and the relevance of questions (28/268 commenting respondents, 10.4%). Several spoke workers described burden and coverage as two faces of the same rigidity: “We can’t skip by our choice we have to answer each and each question” (P0649), and “Not all symptoms are included hence we are entering symptoms manually” (P0558). Hub difficulties were led by technical performance, mostly freezing (44/166 commenting respondents, 26.5%), followed by the quality or scope of suggestions (22/166 commenting respondents, 13.3%), interface and display problems (19/166 commenting respondents, 11.4%) and suggestions that were not generated or displayed (14/166 commenting respondents, 8.4%). The comments showed calibrated rather than wholesale reliance: “We won’t depend on system generated DD for provisional diagnosis as it’s not accurate in all circumstances but it might help in think for possibilities” (P0621), and “Diagnosis options are not diagnoses, rather symptoms” (P0114). When the least-liked prompt listed examples, 82 hub respondents simply named a component (49 the differential diagnosis and 41 symptom modification) without describing a problem.

Requested improvements leaned heavily on the prompted examples: 107 of 148 hub and 148 of 206 spoke improvement responses mentioned only image upload, language support or quicker symptom editing. Unprompted requests were more informative. Spoke workers asked for greater availability of hub doctors and shorter waiting times (39/320 commenting respondents, 12.2%), a service operations issue rather than a CDSS one, and hub clinicians asked for prescription, investigation and record functions (21/166 commenting respondents, 12.7%). Table 5 brings the structured and comment evidence together by workflow issue and separates what was observed from what a future evaluation should measure.

**Table 5.** Joint display: observed evidence and proposed future evaluation.

| **Workflow issue** | **Observed structured and comment evidence** | **Interpretation and uncertainty** | **Proposed accountable function** | **Proposed future indicator (numerator / denominator) and balancing measure** |
| --- | --- | --- | --- | --- |
| Suggestions mostly not generated or not displayed (hub) | 11.1% mostly none generated (up to 13.2% with ambiguous answers); intention 61.9% vs 87.6%. Comments: 14 respondents. | Mixes model scope (31 conditions), display or connectivity failure and nonuse; 6 of 10 who never reviewed suggestions chose this option. | CDSS technical lead with platform operations (proposed) | Consultations with $\geq 1$ suggestion displayed / consultations with completed structured entry, from logs; failure reasons coded. Balance: rate of inappropriate suggestions; hub review time. |
| Suggestions perceived as inappropriate or limited (hub) | 7.9% mostly inappropriate; 40.0% appropriate but different; intention 66.7% vs 89.9%. Comments: 22 respondents. | Clinician agreement is not accuracy; small group, wide interval; weaker among respondents who reviewed suggestions. | Clinical quality lead with CDSS clinical team (proposed) | Sampled output-bearing encounters with sufficient information, rated against a defined reference standard; report concordance categories and unauditable records. Balance: acceptance of inappropriate suggestions; display of model scope to clinicians. |

| Workflow issue | Observed structured and comment evidence | Interpretation and uncertainty | Proposed accountable function | Proposed future indicator (numerator / denominator) and balancing measure |
| --- | --- | --- | --- | --- |
| Question quantity miscalibrated in both directions (spoke) | 15.6% too many, 16.6% too few; satisfaction unchanged (-2.7 percentage points). Comments: burden 82, relevance 28 respondents. | Burden is tolerated but rigid; simple complaints attract unnecessary questions. | CDSS product and workflow lead with spoke service lead (proposed) | Pilot of complaint-specific adaptive questioning: median entry time (timestamp-complete tasks); completed required attributes / applicable attributes in audited encounters; abandoned / initiated entries. Balance: missing critical information. |
| Missing complaints in the symptom library (spoke) | Comments: 68 respondents; examples include skin, pain and follow-up complaints. | Coverage gaps push information into free text, which the model may not use. | CDSS content team (proposed) | Complaints entered as manual free text / all complaints entered, from logs, by state and language. Balance: list length and search time. |
| Freezing, display errors and connectivity (platform) | 38.4% of hub respondents reported technical problems; intention unchanged (-2.8 percentage points). Comments: 44 respondents. | Friction not associated with intention but may affect throughput and data completeness. | Platform operations (proposed) | Sessions with freeze or error events / all sessions; successful connections / call attempts. Balance: abandoned consultations. |
| Training and feature awareness (both roles) | Trained: hub 38.4%, spoke 48.2%; want more training: 66.3%, 74.6%; multilingual search awareness 64.4%. | Training was self-reported; associations with acceptance are descriptive only. | Program training lead (proposed) | Providers meeting a competency criterion / providers assessed; verified feature use / eligible opportunities. Balance: inappropriate use; task time. |

## Discussion

### Principal Findings

Hub clinicians and spoke workers both accepted eSanjeevani’s CDSS, but they accepted different things, and their acceptance moved with different difficulties. For hub clinicians, the experience most closely associated with acceptance was the AI suggestion itself. Intention to keep using the tools fell from 98.4% when suggestions matched their own diagnosis to 66.7% when they were mostly inappropriate and 61.9% when they were mostly absent, whereas the technical problems reported by more than a third of them made almost no difference. For spoke workers, the question burden of structured symptom entry was a common complaint but not a reason for dissatisfaction; satisfaction was essentially flat across the quantity categories. The more useful signal at the spoke was that miscalibration ran in both directions, with similar shares finding the questions too many and too few.

Read together, these findings suggest that hub clinicians do not judge the tool as a whole. They appear to judge it by whether its outputs fit the case in front of them, and they seem to tolerate friction that does not affect that judgment. Two cautions follow. The first is that agreement with the clinician’s own diagnosis is a comfortable standard but not a measure of accuracy. A model that mostly echoes the clinician will be liked, whereas one that offers a correct but unexpected alternative may be rated “appropriate but different” and attract less enthusiasm; in our data, intention among respondents who chose that category was 15.5 percentage points lower than among those whose suggestions matched. Satisfaction therefore cannot be the yardstick for model performance. The second is that “no suggestion generated” is not a single phenomenon. The developers describe an output space of 31 conditions [20], far narrower than the range of complaints seen in primary care, and some clinicians described suggestions that were not displayed or that froze with slow connections. Yet several respondents who reported absent suggestions never reviewed them or were unaware of the feature. Only consultation logs can separate model scope, display failure and nonuse.

At the spoke, the comments explain why burden did not translate into dissatisfaction. Spoke workers valued having options already listed and complained mainly about rigidity: mandatory questions for simple complaints, repeated questions across related complaints and missing common complaints that had to be typed manually. These are calibration problems of a branching instrument rather than objections to structure. Whether this tolerance would hold under heavier caseloads, or partly reflects a habit of answering program surveys favorably, cannot be settled with these data. They matter beyond the spoke, because the structured entry is the input to the hub’s AI suggestions. Burden that leads to skipped or approximate answers, and coverage gaps that push information into free text, would degrade the very data from which suggestions are generated.

### Comparison With Prior Work

Our acceptance levels are broadly consistent with the Odisha evaluation, although its measures differed from ours, in which 94% of community health officers were comfortable with the symptom checker and 90% of hub and sub-hub doctors found the probable-diagnosis feature useful; that evaluation’s interviews also noted that the symptom list lacked options for less common complaints [26]. Earlier eSanjeevani studies described platform-level barriers, such as specialist availability, connectivity and waiting time, among community health officers [23] and doctors [24], and a hub-level comparison of platform versions flagged the need to refine the chief-complaint section [25]. Our spoke workers raised clinician availability unprompted as well, which locates part of their dissatisfaction in service operations rather than in the CDSS. Beyond this shared picture, the present study adds a function-by-role view: which part of the CDSS each role meets, how that part is experienced, and which specific difficulties are associated with acceptance and which are not.

Internationally, health professionals using symptom checkers for triage in Finland reported both streamlined work and new clarification tasks [27], and acceptance of AI in hospitals has been linked to workflow integration and training [35]. The dependence of hub intention on suggestion quality fits experimental evidence that clinicians are helped by good AI support and misled by faulty support [10,11], and that automation bias increases with cognitive load and verification difficulty [36], conditions that high-volume teleconsultation can create. The Kenyan experience is the closest parallel and the most sobering. Clinicians there valued an embedded consultation aid almost universally [12], yet harmful recommendations were sometimes carried into the record [13], and the trial showed no clear clinical benefit [14]. eSanjeevani’s model is a narrower classifier than a language model, but the same caution applies. High acceptance among users is a reason to measure performance carefully, not a substitute for doing so.

### Implications for Future Program Evaluation

Table 5 separates what the survey observed from what should be measured next, and assigns each issue to the function that could act on it. Three priorities stand out. First, the availability of suggestions should be monitored directly from logs, as the share of eligible consultations in which suggestions were generated and displayed, with failures attributed to model scope, integration or connectivity. Second, suggestion quality needs an independent reference. This means sampled encounter audits against a defined diagnostic standard, reported alongside the rate at which clinicians accept inappropriate suggestions, and paired with clear communication to clinicians of the model’s scope [38]. Early-stage human factors evaluation of this kind is what the DECIDE-AI guideline was designed to report [39], and recurring local validation is preferable to a one-time external check [18,19]. Third, the spoke questionnaire should become adaptive and complaint specific, with the share of complaints entered as manual free text tracked as a routine signal of library gaps. Any reduction in questions should be tested against the completeness of clinically required attributes, because shorter entry is only an improvement if critical information survives. Documentation burden itself lacks validated measures [40], and dual data entry is already a recognized workload problem for India’s primary care workers [41].

The remaining issues were largely outside the CDSS. Freezing, connectivity and call routing belong to platform operations, and clinician availability belongs to service delivery; digital tools are not a substitute for functioning health systems [42]. Training is a shared responsibility. Only about half of spoke workers and fewer than half of hub clinicians reported any training, awareness of multilingual search was incomplete, and most respondents in both roles asked for more training.

### Strengths and Limitations

This study has several strengths. It used two instruments designed around the task each role performs, drew respondents from a national program across many states, derived the cohort transparently, prespecified within-role contrasts reported as absolute differences, and tested its main estimates against alternative cohorts, ambiguous answers, possible scale misreading and geographic concentration. It also separated prompted echoes from spontaneous comments, a distinction that changes how the requests for image upload and language support should be read.

The limitations of this study include: All measures are self-reported and were not linked to consultation logs, encounters or outcomes, so the study cannot speak to diagnostic accuracy, consultation time or patient benefit, and the CDSS version at each site was not verified. Participation was roughly one in five eligible providers, submissions clustered after coordinated reminders, and responses were concentrated in a few states; a single district contributed about a fifth of spoke responses. Because the survey was distributed through program channels, favorable responses may be overrepresented. Identity was established by account rather than person, and the hub cohort included some nurses and community health officers whose role on the hub form is uncertain, although restricting to medically qualified respondents did not change the findings. Some respondents appear to have misread the satisfaction scale. The confidence intervals assume independent respondents and do not describe national uncertainty.

## Conclusions

In India’s national telemedicine service, CDSS functions were widely accepted by both the spoke workers who enter structured symptoms and the hub clinicians who review AI suggestions. Hub acceptance was associated with whether suggestions were available and fitted the case rather than with technical friction, and spoke acceptance coexisted with a question burden whose miscalibration ran in both directions. These are reports of experience, not evidence of clinical effect. Their value lies in pointing the next evaluation cycle toward log-based measures of suggestion availability, independent audits of suggestion quality and tests of adaptive symptom questioning, an approach that other assisted telemedicine programs adding AI decision support could adapt to their own workflows.

## Declaration

### Authors’ Contributions

**PB, VG, SK & SS:** Conceptualized the study, Writing – review & editing; **SR:** Data curation, Formal analysis, Methodology, Writing – original draft; **PB:** Writing – original draft; **VG, SK & HS:** Development of survey questionnaire, study protocol, survey coordination, interpretation and Writing – review & editing; **SS, TD, AC:** administered and analyzed the online questionnaire, Writing – review & editing; **KNK:** Writing – review & editing; **HS:** Conceptualization, Methodology, Project administration, Supervision, Validation, Writing – review & editing. All authors reviewed and approved the final manuscript.

### Consent for Publication

Not applicable

### Ethics and Consent

This study was approved by the Ethics Committee of the ICMR Central Ethics Committee on Human Research (CECHR) with reference no. CECHR 002/2025.

## Supporting information

no link available

## Acknowledgments

We sincerely thank ICMR-NIRDH, the hub clinicians, community health officers and other spoke providers who participated in this study. We also thank to the state program coordinators for facilitating survey dissemination and to the eSanjeevani teams at the Ministry of Health and Family Welfare and the Centre for Development of Advanced Computing (C-DAC), Mohali, for their support. We are grateful to all contributors whose efforts and collaboration made this research and its publication possible. Generative AI tools were used solely to improve grammatical accuracy and language clarity. The authors carefully reviewed and verified all content and take full responsibility for the final manuscript.

## Data Availability

Individual-level survey data contain account email addresses and free-text comments and are held by the CDAC, Mohali and the Wadhwani Foundation. Deidentified data may be shared on reasonable request to the corresponding author, subject to approval by the Indian Council of Medical Research and the eSanjeevani program. Aggregate tables are provided in Multimedia Appendix 2.

## Abbreviations

AAM: Ayushman Arogya Mandir
AI: artificial intelligence
AYUSH: Ayurveda, Yoga and Naturopathy, Unani, Siddha and Homoeopathy
C-DAC: Centre for Development of Advanced Computing
CDSS: clinical decision support system
CECHR: Central Ethics Committee on Human Research
CHERRIES: Checklist for Reporting Results of Internet E-Surveys
ICMR: Indian Council of Medical Research
OR: odds ratio
STROBE: Strengthening the Reporting of Observational Studies in Epidemiology

## References

1. eSanjeevani crosses 50 crore milestone; women account for 57% of utilisation. DD India. October 6, 2026. Accessed October 9, 2026. https://ddindia.co.in/2026/10/esanjeevani-crosses-50-crore-milestone-women-account-for-57-of-utilisation/

2. Sood S, Lal K, Bhatia M, et al. Adoption and utilization of India’s eSanjeevani national telemedicine service. Oxf Open Digit Health. 2025;3:oqaf025. doi:10.1093/oodh/oqaf025

3. Dastidar BG, Jani AR, Suri S, Nagaraja VH. Reimagining India’s National Telemedicine Service to improve access to care. Lancet Reg Health Southeast Asia. 2024;30:100480. doi:10.1016/j.lansea.2024.100480

4. Kwan JL, Lo L, Ferguson J, et al. Computerised clinical decision support systems and absolute improvements in care: meta-analysis of controlled clinical trials. BMJ. 2020;370:m3216. doi:10.1136/bmj.m3216

5. Kawamoto K, Houlihan CA, Balas EA, Lobach DF. Improving clinical practice using clinical decision support systems: a systematic review of trials to identify features critical to success. BMJ. 2005;330(7494):765. doi:10.1136/bmj.38398.500764.8F

6. Bright TJ, Wong A, Dhurjati R, et al. Effect of clinical decision-support systems: a systematic review. Ann Intern Med. 2012;157(1):29–43. doi:10.7326/0003-4819-157-1-201207030-00450

7. Agarwal S, Glenton C, Tamrat T, et al. Decision-support tools via mobile devices to improve quality of care in primary healthcare settings. Cochrane Database Syst Rev. 2021;7(7):CD012944. doi:10.1002/14651858.CD012944.pub2

8. Semigran HL, Linder JA, Gidengil C, Mehrotra A. Evaluation of symptom checkers for self diagnosis and triage: audit study. BMJ. 2015;351:h3480. doi:10.1136/bmj.h3480

9. Wallace W, Chan C, Chidambaram S, et al. The diagnostic and triage accuracy of digital and online symptom checker tools: a systematic review. NPJ Digit Med. 2022;5(1):118. doi:10.1038/s41746-022-00667-w

10. Tschandl P, Rinner C, Apalla Z, et al. Human-computer collaboration for skin cancer recognition. Nat Med. 2020;26(8):1229–1234. doi:10.1038/s41591-020-0942-0

11. Jabbour S, Fouhey D, Shepard S, et al. Measuring the impact of AI in the diagnosis of hospitalized patients: a randomized clinical vignette survey study. JAMA. 2023;330(23):2275–2284. doi:10.1001/jama.2023.22295

12. Obong’o C, Njenga G, Otiangala D, et al. Mixed-methods evaluation of clinician experiences and adoption patterns of an EHR-integrated generative AI-based clinical decision support uptake by clinicians in Kenya. BMJ Digit Health AI. 2026;2(1):e000207. doi:10.1136/bmjdhai-2025-000207

13. Agweyu A, Mwaniki P, Musau W, et al. Safety of a large language model-based clinical decision support system in African primary healthcare. Nat Health. 2026;1(6):607–618. doi:10.1038/s44360-026-00082-5

14. Agweyu A, Mwaniki P, Menon V, et al. Generative AI-enabled clinical decision support system in primary care: a pragmatic, cluster-randomized trial. Nat Med. 2026;32(8):3032–3039. doi:10.1038/s41591-026-04503-6

15. Wong A, Otles E, Donnelly JP, et al. External validation of a widely implemented proprietary sepsis prediction model in hospitalized patients. JAMA Intern Med. 2021;181(8):1065–1070. doi:10.1001/jamainternmed.2021.2626

16. Finlayson SG, Subbaswamy A, Singh K, et al. The clinician and dataset shift in artificial intelligence. N Engl J Med. 2021;385(3):283–286. doi:10.1056/NEJMc2104626

17. Kelly CJ, Karthikesalingam A, Suleyman M, Corrado G, King D. Key challenges for delivering clinical impact with artificial intelligence. BMC Med. 2019;17(1):195. doi:10.1186/s12916-019-1426-2

18. Feng J, Phillips RV, Malenica I, et al. Clinical artificial intelligence quality improvement: towards continual monitoring and updating of AI algorithms in healthcare. NPJ Digit Med. 2022;5(1):66. doi:10.1038/s41746-022-00611-y

19. Youssef A, Pencina M, Thakur A, Zhu T, Clifton D, Shah NH. External validation of AI models in health should be replaced with recurring local validation. Nat Med. 2023;29(11):2686–2687. doi:10.1038/s41591-023-02540-z

20. Panda PK, Dutta T, Chander A, et al. AI clinical decision support system (CDSS) for teleconsultations in eSanjeevani, India telemedicine service: a prospective implementation study. medRxiv. Preprint posted online November 25, 2025. doi:10.1101/2025.11.22.25340800

21. Greenhalgh T, Wherton J, Papoutsi C, et al. Beyond adoption: a new framework for theorizing and evaluating nonadoption, abandonment, and challenges to the scale-up, spread, and sustainability of health and care technologies. J Med Internet Res. 2017;19(11):e367. doi:10.2196/jmir.8775

22. Nagaraja VH, Dastidar BG, Suri S, Jani AR. Perspectives and use of telemedicine by doctors in India: a cross-sectional study. Health Policy Technol. 2024;13:100845. doi:10.1016/j.hlpt.2024.100845

23. Vaidya V, Potdar GD, Teli A, Chelluri S. Enhancing rural healthcare delivery in India: eSanjeevani and the community health officer perspective. Natl Med J India. 2026;39(5):306–308. doi:10.25259/NMJI_541_2024

24. Dhinu K, Akoijam BS, Singh KB. Acceptability, benefits, and challenges of teleconsultation: a qualitative study among doctors in Manipur. Indian J Med Res. 2026;164(3):405–411. doi:10.25259/IJMR_614_2026

25. Gupta R, Mandula PP, Dash N, et al. Evolution of India’s National teleconsultation platform e-Sanjeevani: a comparative analysis of versions 2.0 and 1.0, with a preliminary report from the state of Haryana. Health Technol. 2025;15:153–160. doi:10.1007/s12553-024-00907-x

26. Intelehealth. Facility Readiness Assessment and Provider Preparedness of the eSanjeevani – National Telemedicine Service in Odisha, India: midline evaluation (fieldwork April–June 2024). Intelehealth; 2025. Accessed October 9, 2026. https://intelehealth.org/wp-content/uploads/2025/05/FRA_Odisha_Report_Final_23-05-2025.pdf

27. Kujala S, Hörhammer I. Health care professionals’ experiences of web-based symptom checkers for triage: cross-sectional survey study. J Med Internet Res. 2022;24(5):e33505. doi:10.2196/33505

28. Mahlknecht A, Engl A, Piccoliori G, Wiedermann CJ. Supporting primary care through symptom checking artificial intelligence: a study of patient and physician attitudes in Italian general practice. BMC Prim Care. 2023;24(1):174. doi:10.1186/s12875-023-02143-0

29. von Elm E, Altman DG, Egger M, Pocock SJ, Gøtzsche PC, Vandenbroucke JP. The Strengthening the Reporting of Observational Studies in Epidemiology (STROBE) statement: guidelines for reporting observational studies. Lancet. 2007;370(9596):1453–1457. doi:10.1016/S0140-6736(07)61602-X

30. Eysenbach G. Improving the quality of web surveys: the Checklist for Reporting Results of Internet E-Surveys (CHERRIES). J Med Internet Res. 2004;6(3):e34. doi:10.2196/jmir.6.3.e34

31. Wilson EB. Probable inference, the law of succession, and statistical inference. J Am Stat Assoc. 1927;22(158):209–212. doi:10.1080/01621459.1927.10502953

32. Newcombe RG. Interval estimation for the difference between independent proportions: comparison of eleven methods. Stat Med. 1998;17(8):873–890. doi:10.1002/(sici)1097-0258(19980430)17:8<873::aid-sim779>3.0.co;2-i

33. Meade AW, Craig SB. Identifying careless responses in survey data. Psychol Methods. 2012;17(3):437–455. doi:10.1037/a0028085

34. Hsieh HF, Shannon SE. Three approaches to qualitative content analysis. Qual Health Res. 2005;15(9):1277–1288. doi:10.1177/1049732305276687

35. Lambert SI, Madi M, Sopka S, et al. An integrative review on the acceptance of artificial intelligence among healthcare professionals in hospitals. NPJ Digit Med. 2023;6(1):111. doi:10.1038/s41746-023-00852-5

36. Lyell D, Coiera E. Automation bias and verification complexity: a systematic review. J Am Med Inform Assoc. 2017;24(2):423–431. doi:10.1093/jamia/ocw105

38. Sendak MP, Gao M, Brajer N, Balu S. Presenting machine learning model information to clinical end users with model facts labels. NPJ Digit Med. 2020;3:41. doi:10.1038/s41746-020-0253-3

39. Vasey B, Nagendran M, Campbell B, et al. Reporting guideline for the early-stage clinical evaluation of decision support systems driven by artificial intelligence: DECIDE-AI. Nat Med. 2022;28(5):924–933. doi:10.1038/s41591-022-01772-9

40. Moy AJ, Schwartz JM, Chen R, et al. Measurement of clinical documentation burden among physicians and nurses using electronic health records: a scoping review. J Am Med Inform Assoc. 2021;28(5):998–1008. doi:10.1093/jamia/ocaa325

41. Jat M, Gupta M, Prinja S. Implementation status and usability of digital health interventions among health care workers and end users at the primary health care level in Chandigarh, North India: cross-sectional study. JMIR Form Res. 2025;9:e69824. doi:10.2196/69824

42. World Health Organization. WHO Guideline: Recommendations on Digital Interventions for Health System Strengthening. World Health Organization; 2019. Accessed October 9, 2026. https://www.who.int/publications/i/item/978924155

