## Supplementary material for "Provider Experiences with AI Clinical Decision Support in eSanjeevani": no link available

**Multimedia Appendix 1. Survey instruments (field version)**

### Source and scope

The field forms were administered as web forms between 11 to 29 July, 2025. Item wording is reproduced verbatim from the column headers of the export, including the original spelling, capitalization and punctuation. Response options are those offered on the form, listed in their logical order; every single-choice answer in the export matched one of them. Layout, page breaks and help text of the web form are not reproduced.

### Introductory page and consent

Every analyzed record carried the consent statement recorded by the form: “I agree to participate in this feedback survey.” The survey design document specified the following introductory text, reproduced here as written; the line giving contact persons is omitted.

**Purpose (hub form):** “This short survey is designed to gather your feedback on two specific tools integrated into the eSanjeevani platform: (1) the Symptom Summary in the Chief Complaints section (hereafter referred to as Symptom Summary or SS) (2) the System-Suggested Differential Diagnosis (hereafter referred to as Differential Diagnosis or DD). The feedback you provide will help improve these tools and how they function in real-world consultations.”

**Purpose (spoke form):** “This short survey is designed to gather your feedback on the 'Chief Complaints section' & its features. The feedback you provide will help us improve this section and how it functions in real-world consultations.”

**Confidentiality (both forms):** “No personal or patient-identifying data will be collected. All responses will be stored securely and used only for internal insights and potential academic or policy publications in anonymized, aggregate form.”

**Consent (both forms):** “By continuing with this survey, you confirm the following: 1. You are a healthcare provider using the eSanjeevani platform. 2. You are voluntarily choosing to participate. 3. You may skip any question or exit the survey at any time without consequence. 4. Your responses will be anonymized and will not affect your access to or use of eSanjeevani in any way.”

**Table S1. Hub form, field version (35 items)**

| **No.** | **Column** | **Item wording (as exported)** | **Response type** | **Response options offered** | **Use in the analysis** |
| --- | --- | --- | --- | --- | --- |
| 1 | c4 | Age | Background | Free text (number) | Table 1; age cleaning rules (Table S2 in Multimedia Appendix 2) |
| 2 | c5 | Gender | Single choice | Male; Female; Transgender | Table 1 |
| 3 | c6 | Highest Medical Qualification | Background | Free text or list | Table 1 (grouped qualification); role-attribution sensitivity |
| 4 | c7 | Years of Experience in Healthcare | Background | Free text | Table 1; adjustment variable in ordinal models |
| 5 | c8 | State | Background | Free text or list | Table 1; leave-one-state-out sensitivity |
| 6 | c9 | District | Background | Free text or list | Table 1 (concentration by district) |
| 7 | c10 | Are you aware of the symptom summary feature in the chief complaints section? | Single choice | Yes; No | Supporting item |
| 8 | c11 | Are you aware that you can add or modify symptoms in the chief complaints section? | Single choice | Yes; No | Supporting item |
| 9 | c12 | Are you aware of the system suggested differential diagnoses feature? | Single choice | Yes; No | Supporting item; exposure sensitivity (Scenario K) |
| 10 | c13 | How often do you review the symptom summary? | Single choice | Regularly; Occasionally; Never | H1 (review of the symptom summary) |
| 11 | c14 | How often do you review the system suggested differential diagnoses? | Single choice | Regularly; Occasionally; Never | H1 (review of suggested diagnoses); exposure sensitivity (Scenario K) |
| 12 | c15 | How the system suggested differential diagnoses were related to your diagnosis? | Single choice | Always matched my diagnosis; Mostly Clinically appropriate but did not match; Mostly Clinically inappropriate; Mostly No differential diagnosis was generated | H2; problem groups for the inappropriate-suggestion and absent-suggestion contrasts |
| 13 | c16 | The CDSS tools (chief complaint summary & system suggested differential diagnosis) made the duration of the teleconsultation | Single choice | Much Shorter; Slightly Shorter; No Change; Slightly Longer; Much Longer | Supporting item |
| 14 | c17 | How useful are CDSS tools in your work on eSanjeevani? | Single choice | Always Useful; Somewhat Useful; Not Useful | Supporting item; scale-misreading check |
| 15 | c18 | Perceived Usefulness: Please select all statements that reflect your experience using CDSS tools on eSanjeevani | Select all that apply | □ These tools improve the quality of my consultations. □ They help me consider more possible conditions. □ They make me more confident in my decisions. □ They help me arrive at a provisional diagnosis more quickly. □ Overall, these tools are useful on eSanjeevani. | Supporting item |
| 16 | c20 | Perceived Ease of Use: Please select statements which you agree regarding ease of using CDSS tools | Select all that apply | □ The information generated by the tools is clear and easy to understand. □ The tools function effectively across different patient cases. □ Overall, I find the CDSS tools user-friendly. □ It was easy for me to learn how to use the CDSS tools. □ The system supports easy and efficient documentation of prescriptions. | H3 (ease-of-use statements) |
| 17 | c22 | Do you plan to use these CDSS tools features regularly in your clinical practice? | Single choice | Definitely Yes; Probably Yes; Not Sure; Probably No; Definitely No | H4; acceptance measure for hub contrasts (definitely or probably yes) |
| 18 | c23 | What did you like most about these tools? | Open text | Free text | Content analysis: strengths |
| 19 | c24 | What did you like least about these tools? (Symptom modification, differential diagnosis etc) | Open text | Free text | Content analysis: difficulties (prompt listed examples) |
| 20 | c25 | What did you like least about these tools? | Open text | Free text | Content analysis: difficulties |
| 21 | c26 | How satisfied are you with these tools overall? | Single choice | Very Satisfied; Satisfied; Neutral; Dissatisfied; Very dissatisfied | Supporting item; scale-misreading check |
| 22 | c27 | Have you experienced any technical issues while using CDSS tools? | Single choice | Yes; No | Problem group for the technical-problem contrast |
| 23 | c28 | If Yes, please describe the nature of the issue (e.g., freezing, incorrect display, diagnosis not generating, etc.): | Open text | Free text | Content analysis: difficulties |
| 24 | c29 | Any other suggestions or comments: | Open text | Free text | Content analysis: improvements |
| 25 | c30 | Voice-to-Text Prescription Entry: " would you like new feature to record your diagnosis and prescription verbally, and have it auto-transcribed into the relevant sections." | Single choice | Strongly prefer new feature; Somewhat prefer new feature; Neutral; Somewhat prefer current typing method; Strongly prefer current typing method | Supporting item |
| 26 | c31 | Other improvements you'd like to see: (e.g., image uploads, multi-language support, quicker symptom editing) | Open text | Free text | Content analysis: improvements (prompt listed examples) |
| 27 | c32 | Have you undergone any training session on CDSS tools covering Symptom selection, eOPD selection, system suggested differential diagnosis etc. | Single choice | Yes; No | Supporting item (training) |
| 28 | c33 | Would additional training or orientation help in improving your use of CDSS tools in clinical practice? | Single choice | Yes; No | Supporting item (training) |
| 29 | c34 | If yes, what training format would you prefer? (Select all that apply) | Select all that apply | □ Video tutorials □ In-person session □ In-portal/app walk-throughs (on-screen) □ Short printed manual or Job aid □ Live webinar or virtual demonstration □ Other (free text) | Supporting item (training) |
| 30 | c35 | What specific aspects of CDSS tools do you feel need more training or explanation? | Select all that apply | □ Reviewing system Suggested diagnoses □ Modifying chief complaint symptoms □ Modifying symptom attributes in Multiple choice questions □ Prescription documentation □ Other (free text) | Supporting item (training) |
| 31 | c36 | Do you agree with the above all questions for the effectiveness of the questionnaire? | Single choice | Yes, I agree with all questions as they are; No, I have suggestions for changes | Supporting item (feedback on the questionnaire) |
| 32 | c37 | Add (New questions to include): | Open text | Free text | Not analyzed (feedback on the questionnaire) |
| 33 | c38 | Modify (Existing questions to change): Please share the Section Name, Question Number and Suggested Changes | Open text | Free text | Not analyzed (feedback on the questionnaire) |
| 34 | c39 | Remove (Questions to delete): Question Number(s): | Open text | Free text | Not analyzed (feedback on the questionnaire) |
| 35 | c40 | Any other Additional Comments: | Open text | Free text | Not analyzed (feedback on the questionnaire) |

H1-H4: prespecified hub profile items (Table 2 and Figure 3 of the main paper). Supporting items are summarized in Tables S6 and S7 in Multimedia Appendix 2. “Other (free text)”: the export contained a few typed entries alongside the listed options (for example, “Not needed” or “none”); they were not counted as options. Items 19 and 20 share the same stem in the export; item 19 listed examples in parentheses. The form did not route respondents past items that did not apply to them; for example, respondents who never reviewed the suggestions still answered item 12 (Table S15 in Multimedia Appendix 2).

**Table S2. Spoke form, field version (31 items)**

| **No.** | **Column** | **Item wording (as exported)** | **Response type** | **Response options offered** | **Use in the analysis** |
| --- | --- | --- | --- | --- | --- |
| 1 | c4 | Age | Background | Free text (number) | Table 1; age cleaning rules (Table S2 in Multimedia Appendix 2) |
| 2 | c5 | Gender | Single choice | Male; Female; Others | Table 1 |
| 3 | c6 | Highest Medical Qualification | Background | Free text or list | Table 1 (grouped qualification) |
| 4 | c7 | Years of Experience in Healthcare | Background | Free text | Table 1; adjustment variable in ordinal models |
| 5 | c8 | State | Background | Free text or list | Table 1; leave-one-state-out sensitivity |
| 6 | c9 | District | Background | Free text or list | Table 1 (concentration by district) |
| 7 | c10 | Do you know that you can search for symptoms in the Chief Complaints section | Single choice | Yes; No; Not Sure | Supporting item |
| 8 | c11 | Do you know that you can search for symptoms in multiple languages under the Chief Complaints section | Single choice | Yes; No; Not Sure | Supporting item |
| 9 | c12 | Do you know that you can add a symptom manually if it isn't listed in the displayed symptom options. | Single choice | Yes; No; Not Sure | Supporting item |
| 10 | c13 | Do you know that you can upload an image (e.g., for skin conditions) in the Chief Complaints section | Single choice | Yes; No; Not Sure | Supporting item |
| 11 | c14 | How would you rate the quantity of Multiple Choice Questions to be answered for each symptom? (e.g., duration, onset, intensity, etc.) | Single choice | Too many; Appropriate; Too few; I did not notice | S2; problem group and comparator for the question-quantity contrast |
| 12 | c15 | How relevant were the Symptom Questions (duration, onset, intensity etc.) asked in the chief complaint section? | Single choice | Very relevant; Relevant; Not so relevant; Very irrelevant | S3 |
| 13 | c16 | How would you rate the presentation of the symptom summary generated after completing the Chief Complaint section? | Single choice | Clear and well-organized; Clear but could be improved; Somewhat unclear; Not clear at all | S4 |
| 14 | c17 | How useful are CDSS tools in your work on eSanjeevani? | Single choice | Always useful; Somewhat useful; Not useful | Supporting item; scale-misreading check |
| 15 | c18 | Perceived Usefulness: Please select all statements that reflect your experience using CDSS tools on eSanjeevani | Select all that apply | □ These tools improve the quality of my consultations □ They help me consider more possible conditions. □ They make me more confident in my decisions □ They help me arrive at a provisional diagnosis more quickly □ Overall, these tools are useful on eSanjeevani | Supporting item |
| 16 | c19 | Perceived Ease of Use: Please select statements which you agree regarding ease of using CDSS tools | Select all that apply | □ The Chief Complaints section helps me collect and share patient symptoms more effectively. □ It helps me document symptoms more quickly than manual methods. □ It enables me to capture symptoms and attributes more comprehensively, ensuring nothing is missed. □ Overall, the Chief Complaints section is a helpful tool for remote consultations. | S1 (symptom-capture statements); see footnote |
| 17 | c20 | What did you like about the tools such as Chief Complaints section? | Open text | Free text | Content analysis: strengths |
| 18 | c21 | What did you dislike about the Chief Complaints section? | Open text | Free text | Content analysis: difficulties |
| 19 | c22 | How satisfied are you with the chief complaint section overall? | Single choice | Very Satisfied; Satisfied; Neutral; Dissatisfied; Very dissatisfied | S5; acceptance measure for the spoke contrast (satisfied or very satisfied) |
| 20 | c23 | Any other suggestions or comments | Open text | Free text | Content analysis: improvements |
| 21 | c24 | Voice-to-Text Chief Complaint Entry:" Would you like a voice-to-text feature where you speak the patient's chief complaint out loud, and it automatically enters into the chief complaint section (you can review and edit after)" | Single choice | Strongly prefer new feature; Somewhat prefer new feature; Neutral; Somewhat prefer current; Strongly prefer current method | Supporting item |
| 22 | c25 | Other improvements you'd like to see: (e.g., image uploads, multi-language support, quicker symptom editing) | Open text | Free text | Content analysis: improvements (prompt listed examples) |
| 23 | c26 | Have you undergone any training session on CDSS tools as Symptom selection etc. | Single choice | Yes; No | Supporting item (training) |
| 24 | c27 | Would additional training or orientation help improve your use of CDSS tools in clinical practice? | Single choice | Yes; No | Supporting item (training) |
| 25 | c28 | If yes, what training format would you prefer? (Select all that apply) | Select all that apply | □ In-portal/app walk-throughs (on screen) □ In-person session □ Video tutorials □ Live webinar or virtual demonstration □ Short printed manual or Job aid □ Other (free text) | Supporting item (training) |
| 26 | c29 | What specific aspects of CDSS tools do you feel need more training or explanation? | Select all that apply | □ Selecting Speciality OPD □ Selecting symptom attributes in multiple choice questions □ Finding symptoms in drop down □ Entering symptoms □ Other (free text) | Supporting item (training) |
| 27 | c30 | Do you agree with the above questions for the effectiveness of the questionnaire? | Single choice | Yes, I agree with all questions as they are; No, I have suggestions for changes | Supporting item (feedback on the questionnaire) |
| 28 | c31 | Add (New questions to include): | Open text | Free text | Not analyzed (feedback on the questionnaire) |
| 29 | c32 | Modify (Existing questions to change): Please mention Section, Question Number and the suggested change | Open text | Free text | Not analyzed (feedback on the questionnaire) |
| 30 | c33 | Remove (Questions to delete): Question Number(s): | Open text | Free text | Not analyzed (feedback on the questionnaire) |
| 31 | c34 | Any other Additional Comments: | Open text | Free text | Not analyzed (feedback on the questionnaire) |

S1-S5: prespecified spoke profile items (Table 2 and Figure 3 of the main paper). Supporting items are summarized in Tables S6 and S7 in Multimedia Appendix 2.

Item 16: the exported header reads “Perceived Ease of Use”, but its statements concern the usefulness of the chief-complaint section for capturing symptoms. They were analyzed as S1 (symptom-capture statements). The ease-of-use statements for the chief-complaint section appeared only in the pilot version (Table S3).

**Table S3. Differences between the pilot (June 4-9, 2025) and field (July 11-29, 2025) versions**

| **Element** | **Pilot version (excluded)** | **Field version (analyzed)** |
| --- | --- | --- |
| Training items (training received, need for more training, preferred format, aspects needing training) | Absent | Present (hub items 27-30; spoke items 23-26) |
| Hub technical-problem item and its description | Absent | Present (hub items 22-23) |
| Spoke checkbox in column c18 | Ease-of-use statements about the chief-complaint section (“Learning to use the Chief Complaints section was easy for me”, “The interface is clear and intuitive”, “The section is flexible across different patient cases”, “Overall, the Chief Complaints section is easy to use”) | Usefulness statements about the CDSS tools (spoke item 15); no spoke ease-of-use checkbox |
| Hub relation of suggestions to own diagnosis (c15) | “Mostly Not clinically appropriate”; “No differential diagnosis was generated” | “Mostly Clinically inappropriate”; “Mostly No differential diagnosis was generated” |
| Hub ease-of-use statements and voice-to-text response labels | Earlier wording | Reworded |
| Spoke awareness of image upload (c13) | Absent | Present (spoke item 10) |

The pilot submissions were classified as pilot and test entries and excluded from all primary analyses; Scenario C in Table S13 of Multimedia Appendix 2 shows the effect of including them.

**Multimedia Appendix 2. Supplementary methods and results**

***Provider Experiences with AI Clinical Decision Support in eSanjeevani***

*Percentages use respondents who answered as denominators unless stated otherwise. Wilson 95% CIs for proportions and Newcombe hybrid score 95% CIs for differences assume independent respondents. pp: percentage points; OR: odds ratio.*

**Supplementary methods**

Table S1. Retrospective statistical analysis plan: decisions and their timing

Table S2. Variables and cleaning rules

Table S3. Operational definitions of profile items, acceptance measures and contrasts

**Supplementary results**

Table S4. Analysis cohort compared with the excluded pilot wave

Table S5. Combinations ticked on single-response items

Table S6. Supporting single-response items

Table S7. Supporting checkbox items

Table S8. Hub: relation of suggestions to own diagnosis by intention to use

Table S9. Spoke: question quantity by satisfaction

Table S10. Hub proportional-odds model

Table S11. Spoke proportional-odds model

Table S12. Cut-specific logistic models (proportional-odds check)

Table S13. Key estimates under alternative cohorts and codings

Table S14. Leave-one-state-out ranges

Table S15. Measurement checks

Table S16. Codebook for open comments

Table S17. Theme counts by role and domain

**Additional exploratory analysis (not reported in the main paper)**

Table S18. Acceptance by self-reported prior training

### Supplementary methods

**Table S1. Retrospective statistical analysis plan: decisions and their timing**

| **Decision** | **What was decided** | **Timing** | **Reason** |
| --- | --- | --- | --- |
| Analysis unit and cohort | July field wave (July 11-29, 2025); earliest submission per account within each form; accounts that used both forms retained in each role and flagged; no pooling of roles | Specified in the analysis plan before outcome analyses; pilot status of June submissions confirmed by the investigators | Form versions differed; 96 of 97 June records came from five program accounts that used both forms |
| Primary outcome | Role-specific implementation profile (H1-H4, S1-S5) reported as separate item distributions; no composite score | Before outcome analyses | Instruments not psychometrically validated |
| Ambiguous answers | More than one option on a single-response item kept as a separate category; never recoded in primary analyses | Before outcome analyses | Avoids assigning an answer the respondent did not give |
| Secondary outcomes | Four within-role contrasts (hub inappropriate suggestions, absent suggestions, technical problems; spoke too many questions) plus one spoke sensitivity comparator; definitions in Table S3 | Before outcome analyses | Problem and comparator groups locked per contrast, with their own denominators |
| Interval methods | Wilson score intervals for proportions; Newcombe hybrid score intervals for differences | Before outcome analyses | Small problem groups; independent-respondent approximation stated |
| Global tests | Monte Carlo permutation chi-square test (20,000 permutations, seed 1) for the two full-category association questions; Holm adjustment across the two tests | Before association analyses | Sparse cells; one family of two prespecified questions |
| Supporting models | One proportional-odds model per role; adjustment for experience category only; proportional-odds assumption examined with cut-specific logistic models | Before model fitting | Ordinal outcomes grouped into three levels because the lowest categories were sparse |
| Planned sensitivity analyses | Scenarios B-G (cohort definition, ambiguous answers) and I (leave-one-state-out) | Before outcome analyses | Selection, ambiguity and geographic concentration |
| Amendment 1 | Scenario J: hub respondents with medical qualifications only | Added after first results | Some hub-form respondents held nursing or community health qualifications |
| Amendment 2 | Scenario K: hub respondents aware of the suggestions and reviewing them at least occasionally | Added after first results | Skip instructions were not enforced (Table S15) |
| Amendment 3 | Scenario H: exclusion of very dissatisfied responses from respondents who also rated the tools always useful | Added after inspection of distributions | Possible misreading of the satisfaction scale (Table S15) |
| Qualitative analysis | Domains defined by prompt; codebook developed inductively from the full corpus; rule-assisted first pass followed by line-by-line review; prompted echoes tabulated separately | During coding, before theme counts were finalized | Two prompts listed examples |

**Table S2. Variables and cleaning rules**

| **Variable** | **Source format** | **Rule** |
| --- | --- | --- |
| Account (Username) | Email address of the submitting account | Normalized (lowercase, trimmed); replaced with a random study identifier; used only to detect repeat submissions; linkage file held separately |
| Age | Free text | Numeric extraction; dates of birth converted to age at submission; names or nonnumeric entries treated as missing; values <18 or >75 years set to missing |
| Years of experience | Free text | Years and months combined (eg, '4 years 7 months' = 4.58 years); ranges set to their midpoint; word numerals converted; values >45 years or greater than age minus 15 set to missing; grouped as <4, 4-10 and >10 years for the ordinal models |
| Highest qualification | Free text | Cleaned crosswalk from the supplied workbook, then grouped. Hub: MBBS; MBBS with postgraduate degree or diploma; AYUSH; nursing or community health; other or unclear. Spoke: nursing degree; nursing diploma (GNM); AYUSH; community health qualification; MBBS or higher medical; other or unclear |
| State | Free text | Spelling harmonized with the supplied crosswalk |
| Single-response items | Categorical | Original categories retained; more than one option ticked kept as a separate ambiguous category |
| Checkbox items | Semicolon-separated | One indicator per offered option; denominator = respondents answering the item; typed 'Other' entries not counted as options |
| Open text | Free text | Encoding repaired (ftfy); each response classified as empty, placeholder, explicit none or substantive; names typed into demographic fields treated as missing |

Discrepancies with the supplied cleaned worksheet: rows had been re-ordered (records were matched on timestamp and account, with uniqueness asserted); the spoke columns labeled “Perceived Ease of Use” and “Perceived Usefulness” were interchanged relative to the raw export; one hub experience value of “18 months” had been cleaned to 15 years (corrected to 1.5 years); and months had been rounded rather than converted in several experience entries. All analyses were rebuilt from the raw export. AYUSH: Ayurveda, Yoga and Naturopathy, Unani, Siddha and Homoeopathy; GNM: general nursing and midwifery; MBBS: Bachelor of Medicine, Bachelor of Surgery.

**Table S3. Operational definitions of profile items, acceptance measures and contrasts**

| **Component** | **Label** | **Definition** | **Categories or acceptance measure** |
| --- | --- | --- | --- |
| Hub profile | H1 | Review of the symptom summary; review of suggested diagnoses | Regularly; occasionally; never (each item) |
| Hub profile | H2 | Relation of suggestions to own diagnosis | Always matched; mostly appropriate but did not match; mostly inappropriate; mostly not generated; more than one option |
| Hub profile | H3 | Ease-of-use statements | Share endorsing each of five statements |
| Hub profile | H4 | Intention to use the tools regularly | Five categories; affirmative = definitely or probably yes |
| Spoke profile | S1 | Symptom-capture statements about the chief-complaint section | Share endorsing each of four statements |
| Spoke profile | S2 | Quantity of symptom-attribute questions | Too many; appropriate; too few; did not notice |
| Spoke profile | S3 | Relevance of symptom-attribute questions | Very relevant; relevant; not so relevant; very irrelevant |
| Spoke profile | S4 | Presentation of the symptom summary | Clear and well organized; clear but could be improved; somewhat unclear; not clear at all |
| Spoke profile | S5 | Satisfaction with the chief-complaint section | Five categories plus more than one option; favorable = satisfied or very satisfied |
| Hub contrast | Inappropriate suggestions | Problem: mostly inappropriate. Comparator: always matched or mostly appropriate but different (N=153) | Acceptance: affirmative intention (H4) |
| Hub contrast | Absent suggestions | Problem: mostly not generated. Comparator: any unambiguous answer implying suggestions were available, that is, matched, appropriate but different or inappropriate (N=174) | Acceptance: affirmative intention (H4) |
| Hub contrast | Technical problems | Problem: yes. Comparator: no (N=190) | Acceptance: affirmative intention (H4) |
| Spoke contrast | Too many questions | Problem: too many. Comparator: appropriate (N=507) | Acceptance: favorable satisfaction (S5), unambiguous answers |
| Spoke sensitivity | Too many questions | Problem: too many. Comparator: appropriate or too few (N=614) | Acceptance: favorable satisfaction (S5), unambiguous answers |

Each contrast excluded ambiguous answers and categories outside its comparison, so N differs by contrast. Differences are acceptance in the problem group minus acceptance in the comparator. Item wording is given in Tables S1 and S2 in Multimedia Appendix 1.

### Supplementary results

**Table S4. Analysis cohort compared with the excluded pilot wave**

| **Characteristic** | **Analysis cohort (July)** | **Pilot wave, excluded (June)** |
| --- | --- | --- |
| **Hub form** | | |
| Records, n | 190 | 44 |
| Age, median (IQR), years | 33 (29-39) | 36 (31-40) |
| Female, n (%) | 85 (44.7) | 14 (31.8) |
| Qualification: AYUSH degree, n (%) | 8 (4.2) | 0 (0.0) |
| Qualification: MBBS, n (%) | 114 (60.0) | 31 (70.5) |
| Qualification: MBBS with postgraduate qualification, n (%) | 45 (23.7) | 13 (29.5) |
| Qualification: Nursing or community health, n (%) | 21 (11.1) | 0 (0.0) |
| Qualification: Other or unclear, n (%) | 2 (1.1) | 0 (0.0) |
| States represented, n | 13 | 13 |
| **Spoke form** | | |
| Records, n | 662 | 53 |
| Age, median (IQR), years | 30 (27-34) | 32 (29-36) |
| Female, n (%) | 385 (58.2) | 34 (64.2) |
| Qualification: AYUSH degree, n (%) | 109 (16.5) | 8 (15.1) |
| Qualification: Community health qualification, n (%) | 44 (6.6) | 1 (1.9) |
| Qualification: MBBS or higher medical, n (%) | 38 (5.7) | 3 (5.7) |
| Qualification: Nursing degree, n (%) | 374 (56.5) | 27 (50.9) |
| Qualification: Nursing diploma (GNM), n (%) | 73 (11.0) | 7 (13.2) |
| Qualification: Other or unclear, n (%) | 24 (3.6) | 7 (13.2) |
| States represented, n | 15 | 13 |

96 of the 97 pilot records came from five program accounts that used both forms.

**Table S5. Combinations ticked on single-response items**

| **Item** | **Options ticked** | **n** |
| --- | --- | --- |
| Hub: relation of suggestions to own diagnosis | Always matched my diagnosis; Mostly Clinically appropriate but did not match | 9 |
|  | Mostly Clinically appropriate but did not match; Mostly No differential diagnosis was generated | 3 |
|  | Mostly Clinically appropriate but did not match; Mostly Clinically inappropriate | 2 |
|  | Always matched my diagnosis; Mostly Clinically inappropriate | 1 |
|  | Always matched my diagnosis; Mostly No differential diagnosis was generated | 1 |
| Spoke: satisfaction with the chief-complaint section | Neutral; Satisfied | 8 |
|  | Very dissatisfied; Very Satisfied | 3 |
|  | Satisfied; Very Satisfied | 3 |
|  | Very dissatisfied; Dissatisfied; Neutral; Satisfied; Very Satisfied | 2 |
|  | Neutral; Satisfied; Very Satisfied | 2 |
|  | Dissatisfied; Neutral | 1 |
| Hub: effect on consultation duration | Slightly Shorter; No Change | 3 |
|  | Slightly Shorter; Slightly Longer | 1 |
|  | Much Shorter; Slightly Longer | 1 |

These answers were kept as a separate ambiguous category. Scenarios E, F and G in Table S13 bound their effect.

**Table S6. Supporting single-response items**

| **Item** | **Response** | **n/N** | **% (95% CI)** |
| --- | --- | --- | --- |
| Hub: aware of symptom summary | Yes | 180/190 | 94.7 (90.6-97.1) |
|  | No | 10/190 | 5.3 (2.9-9.4) |
| Hub: aware of adding or modifying symptoms | Yes | 162/190 | 85.3 (79.5-89.6) |
|  | No | 28/190 | 14.7 (10.4-20.5) |
| Hub: aware of suggested differential diagnoses | Yes | 177/190 | 93.2 (88.6-96.0) |
|  | No | 13/190 | 6.8 (4.0-11.4) |
| Hub: effect on consultation duration | Much Shorter | 48/190 | 25.3 (19.6-31.9) |
|  | Slightly Shorter | 76/190 | 40.0 (33.3-47.1) |
|  | No Change | 45/190 | 23.7 (18.2-30.2) |
|  | Slightly Longer | 13/190 | 6.8 (4.0-11.4) |
|  | Much Longer | 3/190 | 1.6 (0.5-4.5) |
|  | Multiple selections | 5/190 | 2.6 (1.1-6.0) |
| Hub: usefulness of CDSS tools | Always Useful | 114/190 | 60.0 (52.9-66.7) |
|  | Somewhat Useful | 67/190 | 35.3 (28.8-42.3) |
|  | Not Useful | 9/190 | 4.7 (2.5-8.8) |
| Hub: overall satisfaction with the tools | Very Satisfied | 24/190 | 12.6 (8.6-18.1) |
|  | Satisfied | 109/190 | 57.4 (50.3-64.2) |
|  | Neutral | 43/190 | 22.6 (17.3-29.1) |
|  | Dissatisfied | 5/190 | 2.6 (1.1-6.0) |
|  | Very dissatisfied | 9/190 | 4.7 (2.5-8.8) |
| Hub: any technical problem | Yes | 73/190 | 38.4 (31.8-45.5) |
|  | No | 117/190 | 61.6 (54.5-68.2) |
| Hub: prior training | Yes | 73/190 | 38.4 (31.8-45.5) |
|  | No | 117/190 | 61.6 (54.5-68.2) |
| Hub: additional training would help | Yes | 126/190 | 66.3 (59.3-72.7) |
|  | No | 64/190 | 33.7 (27.3-40.7) |
| Hub: voice-to-text for diagnosis and prescription | Strongly prefer new feature | 22/190 | 11.6 (7.8-16.9) |
|  | Somewhat prefer new feature | 23/190 | 12.1 (8.2-17.5) |
|  | Neutral | 48/190 | 25.3 (19.6-31.9) |
|  | Somewhat prefer current typing method | 46/190 | 24.2 (18.7-30.8) |
|  | Strongly prefer current typing method | 51/190 | 26.8 (21.0-33.6) |
| Hub: agreed with the questionnaire as it is | Yes, I agree with all questions as they are | 185/190 | 97.4 (94.0-98.9) |
|  | No, I have suggestions for changes | 5/190 | 2.6 (1.1-6.0) |
| Spoke: aware of symptom search | Yes | 622/662 | 94.0 (91.9-95.5) |
|  | No | 28/662 | 4.2 (2.9-6.0) |
|  | Not Sure | 12/662 | 1.8 (1.0-3.1) |
| Spoke: aware of multilingual search | Yes | 426/662 | 64.4 (60.6-67.9) |
|  | No | 157/662 | 23.7 (20.6-27.1) |
|  | Not Sure | 79/662 | 11.9 (9.7-14.6) |
| Spoke: aware of manual symptom addition | Yes | 570/662 | 86.1 (83.3-88.5) |
|  | No | 61/662 | 9.2 (7.2-11.7) |
|  | Not Sure | 31/662 | 4.7 (3.3-6.6) |
| Spoke: aware of image upload | Yes | 516/660 | 78.2 (74.9-81.2) |
|  | No | 112/660 | 17.0 (14.3-20.0) |
|  | Not Sure | 32/660 | 4.8 (3.5-6.8) |
| Spoke: usefulness of CDSS tools | Always useful | 480/662 | 72.5 (69.0-75.8) |
|  | Somewhat useful | 162/662 | 24.5 (21.3-27.9) |
|  | Not useful | 20/662 | 3.0 (2.0-4.6) |
| Spoke: prior training | Yes | 319/662 | 48.2 (44.4-52.0) |
|  | No | 343/662 | 51.8 (48.0-55.6) |
| Spoke: additional training would help | Yes | 494/662 | 74.6 (71.2-77.8) |
|  | No | 168/662 | 25.4 (22.2-28.8) |
| Spoke: voice-to-text for chief complaint | Strongly prefer new feature | 58/662 | 8.8 (6.8-11.2) |
|  | Somewhat prefer new feature | 138/662 | 20.8 (17.9-24.1) |
|  | Neutral | 197/662 | 29.8 (26.4-33.3) |
|  | Somewhat prefer current | 101/662 | 15.3 (12.7-18.2) |
|  | Strongly prefer current method | 168/662 | 25.4 (22.2-28.8) |
| Spoke: agreed with the questionnaire as it is | Yes, I agree with all questions as they are | 630/662 | 95.2 (93.3-96.6) |
|  | No, I have suggestions for changes | 32/662 | 4.8 (3.4-6.7) |

N is the number of respondents answering the item. Spoke image-upload awareness was unanswered by 2 respondents. “Multiple selections”: more than one option ticked (Table S5).

**Table S7. Supporting checkbox items**

| **Item** | **Statement or option** | **n/N** | **% (95% CI)** |
| --- | --- | --- | --- |
| Hub: usefulness statements | These tools improve the quality of my consultations. | 104/190 | 54.7 (47.6-61.7) |
|  | They help me consider more possible conditions. | 105/190 | 55.3 (48.2-62.2) |
|  | They make me more confident in my decisions. | 66/190 | 34.7 (28.3-41.7) |
|  | They help me arrive at a provisional diagnosis more quickly. | 78/190 | 41.1 (34.3-48.2) |
|  | Overall, these tools are useful on eSanjeevani. | 106/190 | 55.8 (48.7-62.7) |
| Spoke: usefulness statements | These tools improve the quality of my consultations | 396/662 | 59.8 (56.0-63.5) |
|  | They help me consider more possible conditions. | 313/662 | 47.3 (43.5-51.1) |
|  | They make me more confident in my decisions | 235/662 | 35.5 (31.9-39.2) |
|  | They help me arrive at a provisional diagnosis more quickly | 242/662 | 36.6 (33.0-40.3) |
|  | Overall, these tools are useful on eSanjeevani | 342/662 | 51.7 (47.9-55.4) |
| Hub: preferred training format | Video tutorials | 70/124 | 56.5 (47.7-64.9) |
|  | Live webinar or virtual demonstration | 40/124 | 32.3 (24.7-40.9) |
|  | In-person session | 34/124 | 27.4 (20.3-35.9) |
|  | Short printed manual or Job aid | 27/124 | 21.8 (15.4-29.8) |
|  | In-portal/app walk-throughs (on-screen) | 18/124 | 14.5 (9.4-21.8) |
| Spoke: preferred training format | Video tutorials | 209/477 | 43.8 (39.4-48.3) |
|  | Live webinar or virtual demonstration | 186/477 | 39.0 (34.7-43.4) |
|  | In-person session | 133/477 | 27.9 (24.0-32.1) |
|  | Short printed manual or Job aid | 85/477 | 17.8 (14.6-21.5) |
|  | In-portal/app walk-throughs (on screen) | 119/477 | 24.9 (21.3-29.0) |
| Hub: aspects needing training | Modifying chief complaint symptoms | 87/190 | 45.8 (38.9-52.9) |
|  | Modifying symptom attributes in Multiple choice questions | 77/190 | 40.5 (33.8-47.6) |
|  | Reviewing system Suggested diagnoses | 68/190 | 35.8 (29.3-42.8) |
|  | Prescription documentation | 56/190 | 29.5 (23.4-36.3) |
| Spoke: aspects needing training | Selecting symptom attributes in multiple choice questions | 252/661 | 38.1 (34.5-41.9) |
|  | Finding symptoms in drop down | 218/661 | 33.0 (29.5-36.7) |
|  | Selecting Speciality OPD | 211/661 | 31.9 (28.5-35.6) |
|  | Entering symptoms | 202/661 | 30.6 (27.2-34.2) |

Percentages are shares of respondents endorsing each option and can sum to more than 100%. Training format: denominator = respondents who said additional training would help and answered the format item (hub 124 of 126; spoke 477 of 494). Typed 'Other' entries (mostly 'no' or 'nothing') were retained in the denominator but not counted as options.

**Table S8. Hub: relation of suggestions to own diagnosis by intention to use**

| **Relation of suggestions** | **Definitely Yes** | **Probably Yes** | **Not Sure** | **Probably No** | **Definitely No** | **Total** |
| --- | --- | --- | --- | --- | --- | --- |
| Always matched my diagnosis | 45 | 16 | 0 | 1 | 0 | 62 |
| Mostly Clinically appropriate but did not match | 30 | 33 | 11 | 1 | 1 | 76 |
| Mostly Clinically inappropriate | 4 | 6 | 4 | 1 | 0 | 15 |
| Mostly No differential diagnosis was generated | 7 | 6 | 6 | 2 | 0 | 21 |
| Multiple selections | 9 | 6 | 1 | 0 | 0 | 16 |
| Total | 95 | 67 | 22 | 5 | 1 | 190 |

Global test on unambiguous answers (excluding the multiple-selection row): Monte Carlo permutation chi-square 35.8, n=174, unadjusted P<.001, Holm-adjusted P=.002 (20,000 permutations, seed 1).

**Table S9. Spoke: question quantity by satisfaction with the chief-complaint section**

| **Quantity of questions** | **Very Satisfied** | **Satisfied** | **Neutral** | **Dissatisfied** | **Very dissatisfied** | **Multiple selections** | **Total** |
| --- | --- | --- | --- | --- | --- | --- | --- |
| Appropriate | 61 | 255 | 70 | 4 | 18 | 12 | 420 |
| Too many | 17 | 57 | 22 | 1 | 2 | 4 | 103 |
| Too few | 17 | 57 | 22 | 2 | 9 | 3 | 110 |
| I did not notice | 4 | 16 | 6 | 1 | 2 | 0 | 29 |
| Total | 99 | 385 | 120 | 8 | 31 | 19 | 662 |

Global test on unambiguous answers (rows) and excluding the multiple-selection column: Monte Carlo permutation chi-square 9.81, n=643, unadjusted P=.62, Holm-adjusted P=.62 (20,000 permutations, seed 1).

**Table S10. Hub proportional-odds model: intention to use (n=173)**

| **Term** | **Unadjusted OR (95% CI)** | **Adjusted OR (95% CI)** |
| --- | --- | --- |
| Appropriate but different (vs matched) | 0.23 (0.12-0.47) | 0.23 (0.11-0.46) |
| Mostly inappropriate (vs matched) | 0.11 (0.04-0.35) | 0.10 (0.03-0.33) |
| Mostly not generated (vs matched) | 0.12 (0.04-0.33) | 0.12 (0.04-0.32) |
| Experience <4 years (vs 4-10) | - | 0.64 (0.32-1.29) |
| Experience >10 years (vs 4-10) | - | 0.86 (0.40-1.84) |

Outcome levels: not affirmative < probably yes < definitely yes. ORs below 1 indicate lower odds of a higher outcome level. Respondents with unambiguous exposure answers and a usable experience value; 1 respondent lacked a usable experience value and was excluded from both models.

**Table S11. Spoke proportional-odds model: satisfaction with the chief-complaint section (n=638)**

| **Term** | **Unadjusted OR (95% CI)** | **Adjusted OR (95% CI)** |
| --- | --- | --- |
| Too many (vs appropriate) | 0.98 (0.63-1.52) | 0.93 (0.60-1.46) |
| Too few (vs appropriate) | 0.80 (0.52-1.22) | 0.85 (0.55-1.31) |
| Did not notice (vs appropriate) | 0.57 (0.27-1.21) | 0.53 (0.25-1.14) |
| Experience <4 years (vs 4-10) | - | 0.69 (0.49-0.96) |
| Experience >10 years (vs 4-10) | - | 1.09 (0.67-1.78) |

Outcome levels: not favorable < satisfied < very satisfied. ORs below 1 indicate lower odds of a higher outcome level. Respondents with unambiguous exposure answers and a usable experience value; 5 respondents lacked a usable experience value and were excluded from both models.

**Table S12. Cut-specific logistic models (proportional-odds check, adjusted for experience)**

| **Role** | **Threshold (events/n)** | **Exposure term** | **OR (95% CI)** |
| --- | --- | --- | --- |
| Hub | Affirmative vs not affirmative (146/173) | Appropriate but different (vs matched) | 0.08 (0.01-0.61) |
|  |  | Mostly inappropriate (vs matched) | 0.03 (<0.01-0.30) |
|  |  | Mostly not generated (vs matched) | 0.03 (<0.01-0.22) |
|  | Definitely yes vs lower (85/173) | Appropriate but different (vs matched) | 0.23 (0.11-0.47) |
|  |  | Mostly inappropriate (vs matched) | 0.12 (0.03-0.44) |
|  |  | Mostly not generated (vs matched) | 0.18 (0.06-0.53) |
| Spoke | Favorable vs not favorable (480/638) | Too many (vs appropriate) | 0.81 (0.49-1.36) |
|  |  | Too few (vs appropriate) | 0.74 (0.45-1.19) |
|  |  | Did not notice (vs appropriate) | 0.55 (0.24-1.28) |
|  | Very satisfied vs lower (97/638) | Too many (vs appropriate) | 1.15 (0.64-2.09) |
|  |  | Too few (vs appropriate) | 1.12 (0.62-2.02) |
|  |  | Did not notice (vs appropriate) | 0.44 (0.10-1.90) |

Each row is a separate binary logistic model at one threshold of the ordinal outcome, with the same exposure and experience terms as Tables S10 and S11. Under proportional odds, the ORs at the two thresholds would be similar. Hub estimates at the affirmative threshold rest on few nonaffirmative answers in the reference category (always matched) and are imprecise.

**Table S13. Key estimates under alternative cohorts and codings**

| **Scenario** | **Hub n** | **Hub intention, %** | **Hub inappropriate / absent, %** | **Hub inappropriate contrast, pp (95% CI)** | **Hub absent contrast, pp (95% CI)** | **Hub technical contrast, pp (95% CI)** | **Spoke n** | **Spoke satisfied, %** | **Spoke too many / too few, %** | **Spoke too-many contrast, pp (95% CI)** |
| --- | --- | --- | --- | --- | --- | --- | --- | --- | --- | --- |
| A. Primary cohort | 190 | 85.3 | 7.9 / 11.1 | -23.2 (-48.5 to -4.0) | -25.7 (-47.1 to -7.3) | -2.8 (-14.1 to 7.3) | 643 | 75.3 | 15.6 / 16.6 | -2.7 (-12.8 to 6.0) |
| B. All July submissions | 193 | 85.0 | 7.8 / 10.9 | -22.7 (-48.0 to -3.5) | -25.3 (-46.7 to -6.9) | -3.8 (-15.1 to 6.3) | 647 | 75.3 | 15.6 / 16.7 | -2.3 (-12.4 to 6.3) |
| C. Including the pilot wave | 237 | 85.7 | 10.1 / 10.5 | -15.7 (-35.9 to -1.7) | -32.8 (-52.1 to -14.7) | -3.8 (-15.1 to 6.3) | 699 | 75.3 | 18.5 / 15.4 | -3.9 (-12.9 to 4.1) |
| D. Excluding accounts that used both forms | 180 | 85.6 | 7.2 / 11.1 | -20.9 (-48.1 to -1.8) | -28.3 (-50.0 to -9.1) | -0.1 (-11.6 to 10.0) | 634 | 75.1 | 15.5 / 16.4 | -3.0 (-13.2 to 5.8) |
| E. Ambiguous satisfaction counted unfavorable | - | - | - | - | - | - | 662 | 73.6 | 15.6 / 16.6 | -3.9 (-14.0 to 5.0) |
| F. Ambiguous satisfaction counted favorable if any favorable option | - | - | - | - | - | - | 662 | 75.8 | 15.6 / 16.6 | -3.3 (-13.2 to 5.2) |
| H. Excluding very dissatisfied + always useful (spoke) | - | - | - | - | - | - | 619 | 78.2 | 15.8 / 16.3 | -3.9 (-14.0 to 4.6) |
| J. Hub respondents with medical qualifications only | 167 | 86.2 | 7.8 / 10.8 | -21.0 (-48.2 to -1.8) | -21.6 (-44.9 to -3.3) | -3.7 (-15.9 to 6.7) | - | - | - | - |
| K. Hub respondents aware of and reviewing suggestions | 175 | 88.0 | 6.9 / 8.6 | -16.7 (-45.2 to 0.4) | -30.3 (-54.9 to -9.3) | -2.0 (-13.1 to 7.6) | - | - | - | - |

Hub intention: definitely or probably yes. Spoke satisfied: satisfied or very satisfied. Contrasts are acceptance in the problem group minus the comparator (Table S3). Spoke n and satisfaction use unambiguous satisfaction answers except in Scenarios E and F, which assign ambiguous answers. Scenario C harmonizes pilot-version labels for the hub diagnostic item; the technical-problem item was absent from the pilot form, so that contrast uses July records only. Scenario G (bounds for ambiguous hub diagnostic answers): mostly inappropriate 7.9%-9.5%; mostly not generated 11.1%-13.2%; matched or appropriate 72.6%-77.4%. Scenario I (leave-one-state-out) is shown in Table S14. Scenario ranges are not confidence intervals.

**Table S14. Leave-one-state-out ranges (Scenario I)**

| **Estimate** | **Range across exclusions** |
| --- | --- |
| Hub affirmative intention, % | 83.6 to 86.2 |
| Hub matched or appropriate, % | 70.7 to 73.8 |
| Hub mostly inappropriate, % | 6.6 to 8.6 |
| Hub mostly not generated, % | 10.4 to 12.1 |
| Hub technical problem, % | 37.4 to 39.7 |
| Hub inappropriate contrast, pp | -32.6 to -14.4 |
| Hub absent-suggestion contrast, pp | -29.3 to -22.7 |
| Hub technical contrast, pp | -5.4 to 0.3 |
| Spoke satisfied, % | 74.1 to 77.0 |
| Spoke too many questions, % | 14.9 to 16.7 |
| Spoke too few questions, % | 15.4 to 17.5 |
| Spoke too-many contrast, pp | -4.6 to 1.5 |

Each state with at least five respondents in the role was excluded in turn and the estimates were recomputed; the table gives the lowest and highest values. Point estimates only.

**Table S15. Measurement checks**

| **Check** | **Group** | **n** | **Finding** |
| --- | --- | --- | --- |
| Skip logic not enforced (hub) | Never reviewed the suggested diagnoses | 10 | 6 reported suggestions mostly not generated |
|  | Unaware of the suggested-diagnosis feature | 13 | 5 reported suggestions mostly not generated |
|  | Answered No to all three awareness items | 6 | Answered the later review and relation items nonetheless |
|  | Scenario K: aware and reviewing at least occasionally (n=175) |  | Absent-suggestion contrast -30.3 pp (-54.9 to -9.3); inappropriate-suggestion contrast -16.7 pp (-45.2 to 0.4) |
| Possible scale misreading | Hub: very dissatisfied with the tools | 9 | 8 also rated the tools always useful; 8 intended to keep using them |
|  | Spoke: very dissatisfied with the chief-complaint section | 31 | 24 also rated the tools always useful |
|  | Scenario H: spoke cohort excluding these 24 responses (n=619) |  | Satisfaction 78.2%; too-many contrast -3.9 pp (-14.0 to 4.6) |

pp: percentage points (95% CI). Very dissatisfied answers paired with the most favorable usefulness rating and few critical comments suggest that some respondents read the satisfaction scale in reverse.

**Table S16. Codebook for open comments**

| **Theme** | **Definition** | **Responsible function** |
| --- | --- | --- |
| **Strengths (liked prompts)** | | |
| Ease of use | Ease of use, simplicity, user-friendliness | Strength |
| Structured and comprehensive symptom capture | Structured, comprehensive symptom capture; listed options; search and auto-suggest | Strength |
| Clear summary and presentation | Clear, organized summary or presentation of case information | Strength |
| Diagnostic and decision support | Support for diagnosis, differential diagnosis or decisions | Strength |
| Time saving and speed | Time saving and speed | Strength |
| Access for remote patients | Access for remote patients | Strength |
| **Difficulties and requested improvements** | | |
| Question burden and time to complete | Too many, repeated or mandatory questions; length and time to complete entry | CDSS design (symptom-capture workflow) |
| Symptom or condition coverage gaps | Missing symptoms, complaints or conditions; need to add them manually | CDSS content (symptom library) |
| Relevance and specificity of questions or suggestions | Irrelevant, unnecessary or nonspecific questions or suggestions | CDSS content (question logic) |
| Symptom modification and editing | Difficulty modifying or editing symptoms; requests for quicker editing | CDSS interface |
| Quality or scope of diagnostic suggestions | Suggested diagnoses inaccurate, mismatched, nonspecific or limited in range | CDSS model (suggestion quality or scope) |
| Diagnostic suggestions not generated or not displayed | Suggested diagnoses not generated or not displayed | CDSS model or integration (suggestion availability) |
| Interface, navigation and display | Navigation, display, clicks, fields, interface layout | Interface (CDSS or platform) |
| Technical performance (freezing, lag, errors) | Freezing, lag, slow loading, errors, logout, bugs | Platform performance |
| Connectivity, audio-video and call handling | Network, audio-video, call connection, overlapping calls | Platform connectivity |
| Clinician availability, waiting and routing | Clinician availability, waiting time, hub routing and scheduling | Service operations |
| Image upload and quality | Image upload, quality and handling | Platform feature |
| Language support | Regional or multiple language support | Platform or CDSS feature |
| Voice input | Voice input or dictation | Platform feature |
| Prescription, investigation and record functions | Prescription, drug list, investigations, vitals, records, follow-up | Platform clinical functions |
| Training and information quality at entry | Training needs and quality of information entered at the spoke | Training and data quality at entry |

Responses were first classified as empty, placeholder, explicit none or substantive. Difficulty and improvement themes share one codebook because the same issue could be reported as a problem or as a request. A response could receive several codes; each respondent was counted at most once per theme within a domain.

**Table S17. Theme counts by role and domain**

| **Theme** | **Respondents, n** | **% of commenters** | **% of cohort** |
| --- | --- | --- | --- |
| **Hub: strengths (respondents commenting: 176)** | | | |
| Diagnostic and decision support | 63 | 35.8 | 33.2 |
| Ease of use | 48 | 27.3 | 25.3 |
| Structured and comprehensive symptom capture | 37 | 21.0 | 19.5 |
| Time saving and speed | 23 | 13.1 | 12.1 |
| Clear summary and presentation | 10 | 5.7 | 5.3 |
| Access for remote patients | 2 | 1.1 | 1.1 |
| Substantive but no specific theme (general comments) | 29 | - | - |
| **Hub: difficulties (respondents commenting: 166)** | | | |
| Technical performance (freezing, lag, errors) | 44 | 26.5 | 23.2 |
| Quality or scope of diagnostic suggestions | 22 | 13.3 | 11.6 |
| Interface, navigation and display | 19 | 11.4 | 10.0 |
| Question burden and time to complete | 14 | 8.4 | 7.4 |
| Diagnostic suggestions not generated or not displayed | 14 | 8.4 | 7.4 |
| Relevance and specificity of questions or suggestions | 13 | 7.8 | 6.8 |
| Prescription, investigation and record functions | 12 | 7.2 | 6.3 |
| Connectivity, audio-video and call handling | 11 | 6.6 | 5.8 |
| Clinician availability, waiting and routing | 9 | 5.4 | 4.7 |
| Symptom or condition coverage gaps | 5 | 3.0 | 2.6 |
| Symptom modification and editing | 5 | 3.0 | 2.6 |
| Training and information quality at entry | 3 | 1.8 | 1.6 |
| Image upload and quality | 1 | 0.6 | 0.5 |
| Substantive but no specific theme (general comments) | 18 | - | - |
| **Hub: improvements (respondents commenting: 166)** | | | |
| Image upload and quality | 72 | 43.4 | 37.9 |
| Symptom modification and editing | 35 | 21.1 | 18.4 |
| Prescription, investigation and record functions | 21 | 12.7 | 11.1 |
| Language support | 20 | 12.0 | 10.5 |
| Technical performance (freezing, lag, errors) | 14 | 8.4 | 7.4 |
| Interface, navigation and display | 12 | 7.2 | 6.3 |
| Connectivity, audio-video and call handling | 10 | 6.0 | 5.3 |
| Clinician availability, waiting and routing | 10 | 6.0 | 5.3 |
| Quality or scope of diagnostic suggestions | 9 | 5.4 | 4.7 |
| Training and information quality at entry | 8 | 4.8 | 4.2 |
| Question burden and time to complete | 3 | 1.8 | 1.6 |
| Voice input | 3 | 1.8 | 1.6 |
| Symptom or condition coverage gaps | 2 | 1.2 | 1.1 |
| Relevance and specificity of questions or suggestions | 2 | 1.2 | 1.1 |
| Diagnostic suggestions not generated or not displayed | 1 | 0.6 | 0.5 |
| Substantive but no specific theme (general comments) | 21 | - | - |
| **Spoke: strengths (respondents commenting: 470)** | | | |
| Structured and comprehensive symptom capture | 234 | 49.8 | 35.3 |
| Ease of use | 66 | 14.0 | 10.0 |
| Diagnostic and decision support | 49 | 10.4 | 7.4 |
| Clear summary and presentation | 34 | 7.2 | 5.1 |
| Time saving and speed | 23 | 4.9 | 3.5 |
| Access for remote patients | 3 | 0.6 | 0.5 |
| Substantive but no specific theme (general comments) | 162 | - | - |
| **Spoke: difficulties (respondents commenting: 268)** | | | |
| Question burden and time to complete | 82 | 30.6 | 12.4 |
| Symptom or condition coverage gaps | 68 | 25.4 | 10.3 |
| Relevance and specificity of questions or suggestions | 28 | 10.4 | 4.2 |
| Technical performance (freezing, lag, errors) | 10 | 3.7 | 1.5 |
| Interface, navigation and display | 6 | 2.2 | 0.9 |
| Connectivity, audio-video and call handling | 5 | 1.9 | 0.8 |
| Image upload and quality | 3 | 1.1 | 0.5 |
| Clinician availability, waiting and routing | 3 | 1.1 | 0.5 |
| Language support | 3 | 1.1 | 0.5 |
| Prescription, investigation and record functions | 2 | 0.7 | 0.3 |
| Quality or scope of diagnostic suggestions | 1 | 0.4 | 0.2 |
| Substantive but no specific theme (general comments) | 76 | - | - |
| **Spoke: improvements (respondents commenting: 320)** | | | |
| Symptom modification and editing | 73 | 22.8 | 11.0 |
| Image upload and quality | 52 | 16.2 | 7.9 |
| Language support | 39 | 12.2 | 5.9 |
| Clinician availability, waiting and routing | 39 | 12.2 | 5.9 |
| Symptom or condition coverage gaps | 30 | 9.4 | 4.5 |
| Question burden and time to complete | 26 | 8.1 | 3.9 |
| Connectivity, audio-video and call handling | 14 | 4.4 | 2.1 |
| Interface, navigation and display | 14 | 4.4 | 2.1 |
| Technical performance (freezing, lag, errors) | 14 | 4.4 | 2.1 |
| Prescription, investigation and record functions | 11 | 3.4 | 1.7 |
| Relevance and specificity of questions or suggestions | 7 | 2.2 | 1.1 |
| Voice input | 6 | 1.9 | 0.9 |
| Training and information quality at entry | 1 | 0.3 | 0.2 |
| Substantive but no specific theme (general comments) | 68 | - | - |

Cohort: hub n=190; spoke n=662. Respondents with at least one substantive comment: hub 187, spoke 533. Hub respondents who only named a least-liked component without describing a problem: 82 (differential diagnosis 49, symptom modification 41); not counted as difficulties. Improvement responses limited to the prompted examples (image uploads, multilanguage support, quicker symptom editing): hub 107 of 148, spoke 148 of 206.

### Additional exploratory analysis

**Table S18. Acceptance by self-reported prior training**

| **Outcome** | **Trained, n/N (%)** | **Not trained, n/N (%)** | **Difference, pp (95% CI)** |
| --- | --- | --- | --- |
| Hub: affirmative intention | 67/73 (91.8%) | 95/117 (81.2%) | 10.6 (0.1 to 19.7) |
| Spoke: satisfied or very satisfied | 239/309 (77.3%) | 245/334 (73.4%) | 4.0 (-2.7 to 10.6) |

Not prespecified and not reported in the main paper. Unadjusted; training was self-reported and may reflect engagement with the program rather than cause higher acceptance. Spoke analysis uses unambiguous satisfaction answers.

**Multimedia Appendix 3. STROBE checklist for cross-sectional studies**

***Provider Experiences with AI Clinical Decision Support in eSanjeevani***

Checklist items follow the STROBE statement for cross-sectional studies (von Elm et al, Lancet 2007). Recommendations are abbreviated; locations refer to section headings of the main paper and to tables in the multimedia appendices.

| **Section and topic** | **Item** | **Recommendation (abbreviated)** | **Location in the manuscript** |
| --- | --- | --- | --- |
| Title and abstract | 1 | (a) Indicate the study design in the title or abstract. (b) Provide an informative and balanced summary of what was done and what was found | Title; Abstract |
| **Introduction** | | | |
| Background/ rationale | 2 | Explain the scientific background and rationale | Introduction: Background |
| Objectives | 3 | State specific objectives, including any prespecified hypotheses | Introduction: Objective |
| **Methods** | | | |
| Study design | 4 | Present key elements of study design early in the paper | Methods: Study Design and Reporting |
| Setting | 5 | Describe the setting, locations and relevant dates, including periods of recruitment, exposure and data collection | Methods: Setting and the Clinical Decision Support Functions; Participants and Recruitment; Survey Instruments; Figure 1 |
| Participants | 6 | Give the eligibility criteria and the sources and methods of selection of participants | Methods: Participants and Recruitment; Data Processing and Analysis Cohort |
| Variables | 7 | Clearly define all outcomes, exposures, potential confounders and effect modifiers | Methods: Outcomes; Statistical Analysis; Table S3 in Multimedia Appendix 2 |
| Data sources/ measurement | 8 | For each variable of interest, give sources of data and details of methods of assessment | Methods: Survey Instruments; Tables S1 and S2 in Multimedia Appendix 1 |
| Bias | 9 | Describe any efforts to address potential sources of bias | Methods: Statistical Analysis (sensitivity analyses); Tables S13-S15 in Multimedia Appendix 2; Discussion: Strengths and Limitations |
| Study size | 10 | Explain how the study size was arrived at | Methods: Participants and Recruitment (all eligible providers invited; no sample size calculation for this analysis) |
| Quantitative variables | 11 | Explain how quantitative variables were handled in the analyses | Methods: Data Processing and Analysis Cohort; Statistical Analysis; Table S2 in Multimedia Appendix 2 |
| Statistical methods | 12 | (a) Describe all statistical methods, including those used to control for confounding. (b) Methods used to examine subgroups and interactions. (c) How missing data were addressed. (d) If applicable, analytical methods taking account of sampling strategy. (e) Sensitivity analyses | Methods: Statistical Analysis; Tables S1, S12 and S13 in Multimedia Appendix 2. No weighting; intervals assume independent respondents. No subgroup or interaction analyses. No imputation |
| **Results** | | | |
| Participants | 13 | (a) Report numbers of individuals at each stage of the study. (b) Give reasons for nonparticipation at each stage. (c) Consider use of a flow diagram | Results: Participants; Figure 2 |
| Descriptive data | 14 | (a) Give characteristics of study participants. (b) Indicate the number of participants with missing data for each variable of interest | Table 1; Table 2; Tables S4-S7 in Multimedia Appendix 2 |
| Outcome data | 15 | Report numbers of outcome events or summary measures | Table 2; Figure 3 |
| Main results | 16 | (a) Give unadjusted estimates and, if applicable, confounder-adjusted estimates and their precision. (b) Report category boundaries when continuous variables were categorized. (c) If relevant, consider translating estimates of relative risk into absolute risk | Results: Acceptance Alongside Reported Difficulties; Table 3; Figure 4; Tables S8-S12 in Multimedia Appendix 2 |
| Other analyses | 17 | Report other analyses done, for example, analyses of subgroups and interactions, and sensitivity analyses | Results: Sensitivity Analyses and Measurement Checks; Tables S13-S15 in Multimedia Appendix 2 |
| **Discussion** | | | |
| Key results | 18 | Summarize key results with reference to study objectives | Discussion: Principal Findings |
| Limitations | 19 | Discuss limitations, taking into account sources of potential bias or imprecision; discuss direction and magnitude of any potential bias | Discussion: Strengths and Limitations |
| Interpretation | 20 | Give a cautious overall interpretation considering objectives, limitations, multiplicity of analyses, results from similar studies and other relevant evidence | Discussion: Principal Findings; Comparison with Prior Work; Implications for Future Program Evaluation |
| Generalizability | 21 | Discuss the generalizability (external validity) of the study results | Discussion: Strengths and Limitations; Conclusions |
| **Other information** | | | |
| Funding | 22 | Give the source of funding and the role of the funders for the present study | No Funding |

**Multimedia Appendix 4. Checklist for Reporting Results of Internet E-Surveys (CHERRIES)**

***Provider Experiences with AI Clinical Decision Support in eSanjeevani***

Items follow the CHERRIES checklist (Eysenbach, J Med Internet Res 2004). Locations refer to section headings of the main paper and to the multimedia appendices.

| **Item category** | **Checklist item** | **How the item was addressed** | **Location** |
| --- | --- | --- | --- |
| **Design** | | | |
|  | Describe survey design | Cross-sectional web survey using two role-specific forms (hub and spoke), distributed to a protocol-defined multistate eligible population; participation was voluntary, so the respondents were a self-selected subset of those invited. | Methods: Study Design and Reporting; Participants and Recruitment |
| **IRB approval and informed consent process** | | | |
|  | IRB approval | ICMR-CECHR approval of the analysis and publication protocol (CECHR 002/2025). | Methods: Ethical Considerations |
|  | Informed consent | The introductory page described the purpose, voluntariness and use of the data in aggregate publications; consent was given by ticking a statement before the first question. | Methods: Ethical Considerations; Multimedia Appendix 1 |
|  | Data protection | Account email addresses recorded by the form platform were replaced with random identifiers; the linkage file was held separately; only aggregate results are reported. | Methods: Data Processing and Analysis Cohort; Ethical Considerations |
| **Development and pretesting** | | | |
|  | Development and testing | Forms were piloted June 4-9, 2025; pilot entries were excluded; the field version differed from the pilot version. | Methods: Survey Instruments; Table S3 in Multimedia Appendix 1 |
| **Recruitment process and description of the sample having access to the questionnaire** | | | |
|  | Open survey versus closed survey | Closed in intent: links were circulated to eligible providers through official program channels. | Methods: Participants and Recruitment |
|  | Contact mode | Links were circulated by C-DAC state program coordinators through official eSanjeevani communication channels. | Methods: Participants and Recruitment |
|  | Advertising the survey | No public advertising. | Methods: Participants and Recruitment |
| **Survey administration** | | | |
|  | Web/email | Web form; the export format (timestamp and account-email fields) is consistent with Google Forms. | Methods: Participants and Recruitment |
|  | Context | Program feedback survey on eSanjeevani CDSS functions. | Introduction; Methods: Setting and the Clinical Decision Support Functions |
|  | Mandatory/voluntary | Voluntary. | Methods: Participants and Recruitment |
|  | Incentives | No incentives was given participants | Methods: Participants and Recruitment |
|  | Time/date | Pilot June 4-9, 2025 (excluded); field wave July 11-29, 2025. | Methods: Survey Instruments; Results: Participants |
|  | Randomization of items or questionnaires | No. | Not applicable |
|  | Adaptive questioning | No adaptive questioning; the form did not route respondents past items that did not apply to them. | Results: Sensitivity Analyses and Measurement Checks; Table S15 in Multimedia Appendix 2 |
|  | Number of items | Hub field form 35 items; spoke field form 31 items, including open-text and questionnaire-feedback items. | Tables S1 and S2 in Multimedia Appendix 1 |
|  | Completeness check | Closed items were answered by all analyzed respondents, which suggests that responses were required. | Results; Table 2 footnote |
| **Response rates** | | | |
|  | Unique site visitor | Not available (no page-view logs). | Not applicable |
|  | View rate | Not available. | Not applicable |
|  | Participation rate | Responding accounts divided by eligible providers: hub 190/889 (21.4%); spoke 662/3426 (19.3%). Approximate, because respondents could not be linked to the eligibility list. | Results: Participants; Figure 2 |
|  | Completion rate | Not available (partial responses were not recorded by the platform). | Not applicable |
| **Preventing multiple entries from the same individual** | | | |
|  | Cookies used | No. | Not applicable |
|  | IP check | No. | Not applicable |
|  | Log file analysis | Account email addresses were used to identify repeat submissions; the earliest submission was retained (later repeats excluded: hub 3, spoke 4). | Methods: Data Processing and Analysis Cohort; Figure 2 |
|  | Registration | Submissions were linked to the account used to open the form. | Methods: Data Processing and Analysis Cohort |
| **Analysis** | | | |
|  | Handling of incomplete questionnaires | No incomplete questionnaires; multiple selections on single-response items kept as a separate category; no imputation. | Methods: Data Processing and Analysis Cohort; Statistical Analysis; Table S5 in Multimedia Appendix 2 |
|  | Questionnaires submitted with an atypical timestamp | Completion time was not available; submission dates were examined (clustering on July 21-23 reported). | Results: Participants |
|  | Statistical correction | No weighting or propensity adjustment; intervals assume independent respondents; leave-one-state-out sensitivity analyses. | Methods: Statistical Analysis; Table S14 in Multimedia Appendix 2 |
